# Leveraging a Patient-Derived Tumoroid Platform for Precision Radiotherapy: Uncovering DNA Damage Repair Inhibitor-Mediated Radiosensitization and Therapeutic Resistance in Rectal Cancer

**DOI:** 10.1101/2025.01.15.25320629

**Authors:** W Zambare, H Huang, C Wu, S Yoder, Y Gao, J Kim, H Kalvin, M Del Latto, A Garcia, S Meguro, P Bleu, MJ Kim, M Fiasconaro, A Bercz, A Kim, MR Weiser, E Pappou, A Cercek, K Ganesh, PB Paty, J Garcia-Aguilar, M Gonen, X Chen, JJ Smith, PB Romesser

## Abstract

**Background:** Precision radiation strategies that expand the therapeutic window by selectively sensitizing tumors and sparing normal tissues are needed. We developed a matched tumoroid-organoid preclinical platform to identify and characterize personalized radiosensitization strategies.

**Methods:** We established 15 rectal cancer-derived tumoroids and 3 matched normal rectal epithelial organoids. Whole exome sequencing characterized mutation profiles and phylogenetic relationships. Tumoroids were treated with one of four DNA damage repair inhibitors (DDRi; ATMi, DNA-PKi, PARPi, or ATRi), 5-fluorouracil, or a DMSO control, followed by escalating doses of radiation. Cell viability was measured, and intrinsic radiosensitivity as well as radiosensitizer efficacy were characterized using linear regression models. Four tumoroids were derived from one patient at distinct stages and disease sites, including pre-progression tumoroids (primary tumor, splenic metastasis) and post-progression tumoroids (rectal recurrence and vaginal recurrence).

**Results:** Tumoroid radiosensitivity demonstrated variability, paralleling the spectrum of clinical responses seen in rectal cancer. Genomic analyses revealed two distinct mutational signatures (SBS14 and SBS17b) associated with radioresistance. Radiation sensitization by DDRis was highly heterogeneous, depending on both specific tumoroid and inhibitor choice. In the subset of four tumoroids derived pre- and post-progression, post-progression tumoroids demonstrated greater radioresistance and diminished DDRi-induced radiosensitization. Phylogenetic analysis revealed increased clonal and subclonal complexity in these radiation and DDRi-resistant tumoroids. Lastly, comparing matched patient-derived tumoroids and normal organoids established that tumoroids were generally more radiosensitive and exhibited enhanced DDRi-mediated radiosensitization compared to normal organoids. However, the optimal DDRi for maximizing therapeutic index varied among tumoroids.

**Conclusion:** To our knowledge, this is the first *ex vivo* study to systematically quantify intrinsic radiation sensitivity and DDRi sensitization in a comprehensive, patient-specific tumoroid platform. Our findings underscore the utility of a patient-matched tumoroid–organoid model as a platform to quantify and predict personalized responses to radiation and DDR inhibitors. Moreover, by comparing tumoroids from a single patient across different disease stages, our model reveals how tumors adapt to therapeutic pressure and develop increased radioresistance, offering a valuable potential framework for guiding precision radiotherapy. Moreover, the data show that DDRi efficacy is largely tumor-selective and not solely predicted by mutational profiles, highlighting opportunities to refine treatment for radioresistant tumors, minimize injury to normal tissues, and adapt therapy over disease progression.

## Introduction

Radiotherapy (RT) is a cornerstone of cancer therapy, delivered to approximately two-thirds of all cancer patients throughout their disease course, with curative intent in up to 40% of these patients. However, the long-term adverse effects of RT underscore the need for personalized strategies that maintain efficacy while reducing harm to normal tissues.[1] Although the potential of radiation-drug combinations is promising, progress in this area has been limited.[2, 3] Between 2016 and 2021, the US Food and Drug Administration (FDA) approved more than 200 oncology drugs, yet only one – cetuximab – was specifically approved for concomitant treatment with RT.[4] Recognizing this gap, the FDA has identified RT-drug combinations as an unmet clinical need.[5, 6] Despite this, advances have been hampered by the lack of robust human models that accurately mimic tumor and normal tissue biology to assess the therapeutic index of RT-drug combinations.

Recent advances in *ex vivo* organoid/tumoroid cultures offer a promising solution. Organoid/tumoroid cultures recapitulate the three-dimensional architecture, genetic heterogeneity, functional complexity, and even aspects of the tissue/tumor microenvironment more faithfully than traditional cell-lines.[7] Utilizing matched tumor-derived tumoroids and normal epithelial organoids derived from the same patient presents a unique opportunity to simultaneously evaluate tumor-specific radiosensitization and normal tissue toxicity. By leveraging these patient-specific models, we aim to characterize combination strategies that expand the therapeutic window and address the longstanding limitation of normal tissue injury.

Rectal cancer offers a practical and clinically relevant proof of concept for developing such personalized tumoroid-organoid platforms. Radiotherapy is a key treatment modality for rectal cancer, making radiosensitization strategies highly pertinent.[8, 9] Moreover, rectal tumors and adjacent normal epithelium are readily accessible, enabling the establishment of matched tumoroid and organoid cultures.[7] Beyond practicality, a rising incidence of rectal cancer in younger adults creates an urgent clinical need for personalized approaches that improve response rates, reduce toxicity, and increase organ preservation.[10-13] Accordingly, rectal cancer serves as an ideal model for evaluating novel combinations of RT and DNA damage repair inhibitors (DDRis) as radiosensitizers, with findings potentially generalizable to other malignancies.

## Methods

### Patient-derived Rectal Cancer Tumoroids and Normal Epithelial Organoids

With the consent of our patients, biopsies were collected in the clinical setting and cultured into representative tumoroids as part of our growing biorepository (IRB Protocol 16-1071). In this study, “tumoroids” are defined as rectal tumor-derived and “organoids” are defined as normal rectal tissue-derived. The development and validation of our rectal cancer tumoroid platform is described in a previous work.[7] In brief, a biopsy of a rectal tumor is washed thoroughly, manually chopped, and further broken down with digestion buffer. The processed cells are further digested on the warmer, subsequently trypsinized, and then plated within Matrigel domes. With the addition of tumor media, culturing continues under standard conditions yielding the growth of tumoroids. Normal organoids are derived in a similar process with some key modifications and cultured in full media (see Supplementary Methods).

In this study, we highlight fifteen tumoroids derived from eleven patients. Tumoroids were derived from various disease sites including primary tumors, distant metastases, and tumor recurrences. Of these, four tumoroids were derived from the same patient at different disease sites and timepoints. Additionally, two tumoroids were derived at the same timepoint from two heterogeneous areas of the same tumor. Finally, for three patients, adjacent normal rectal tissues were also biopsied and processed into matched normal tissue-derived organoids.

The experimental design represented four different treatment conditions: vehicle control, drug treatment only, radiation only, and combination therapy with drug treatment followed by radiation. Organoids and tumoroids were treated with a vehicle control (DMSO), a chemotherapy control (5-fluorouracil, 5-FU), or one of four DDRi drugs: ATM inhibitor (ATMi), DNA-PK inhibitor (DNA-PKi), PARP inhibitor (PARPi), or ATR inhibitor (ATRi). Inhibitor information is found in Supplementary Table 1. Pilot studies determined appropriate DDRi dosages for optimal sensitization effect without toxicity (see Supplementary Methods). Two hours after drug treatment, the treated organoids and tumoroids were exposed to escalating doses of radiation (0, 2, 4, or 6 Gy) using an X-Rad320 irradiator. They were subsequently incubated for five days and then re-dosed with their corresponding drugs. On day nine, cell viability was measured in each condition using the CellTiter-Glo 2.0 assay (Promega, Catalogue #G9241), following the kit protocol. The experiment was carried out in at least biologic triplicate for each tumoroid or organoid, and each had anywhere from three to four experiments total. The procedure for normal organoids was identical but used complete medium instead of tumor medium, with pilot studies demonstrating that medium choice did not affect drug efficacy. Supplementary Figure 1 displays a schematic of the experiment, with additional information in Supplementary Methods.

Validation of inhibition of several drug targets or downstream targets was achieved by immunohistochemistry. Tumoroids were cultured in all four treatment conditions (no treatment, drug only, radiation only, and combination drug and radiation) and fixed. After paraffin embedding and transfer onto slides, staining was used to evaluate for the effect of inhibitor treatment on downstream targets. Phospho-KAP1 was used to characterize ATM activity, phospho-DNA-PK was used to characterize DNA-PK activity, and phospho-CHK1 was used to characterize ATR activity. Relative protein expression was quantified by the metric of optical density in the software Halo V4.0.

Inhibition of PARP was demonstrated by the expression of cleaved PARP. Tumoroids in all treatment conditions (no treatment, drug only, radiation only, combination drug and radiation treatment) underwent protein extraction using standard RIPA buffer and manual vortex agitation. The Simple Western assay on the Jess Automated Western Blot system (ProteinSimple, Biotechne, Catalogue #004-650) was used evaluate each sample. Protein normalization was performed using an associated module (#DM-PN02, ProteinSimple). The Compass software v6.3.0 (ProteinSimple) was used for protein quantification and blot construction. See Supplementary Methods for additional information and Supplementary Table 2 for a list of antibodies utilized.

### Statistical Analysis

In all modeling performed, cell viability was our outcome of interest. Prior to performing any modeling, cell viability values were standardized by radiation dose 0 Gy, and then log-transformed (base 10) for normality. Several regression models were assessed, and a linear regression model was selected for ease of interpretation and having similar, if not better, model fit to the data as compared to a linear quadratic model. Adjusted R^2^ values were used to check model fit.

To assess the intrinsic tumoroid radiosensitivity in the absence of drug treatment, a linear regression model adjusting for radiation dose and experiment number was created for each tumoroid and organoid using samples exposed to DMSO (vehicle control). The coefficient for radiation dose in this model was used to define percent cell kill.

Another linear regression model was created for each tumoroid and organoid to assess the role of radiosensitivity and drug choice. Model terms included radiation dose, drug (DMSO, 5-FU, or one of the DDRis described above), experiment, and an interaction term to adjust for the relationship between radiation dose and drug. The coefficients from this model were used to calculate the slope, representing the fold change in cell viability as radiation increased, and slope ratio, a comparison of the slope of interest to the slope for DMSO, for each tumoroid/drug combination. The calculated slope from this model was also used to compare tumoroids to matched normal samples (when available). The range in values were illustrated using heatmaps.

Kaplan-Meier methods were used to estimate overall survival (OS) and progression free survival (PFS) by radiosensitivity (categorized as radioresistant or radiosensitive using the median percent cell kill). OS was defined as time from treatment start to death or last follow up and PFS was defined as time from treatment start to progression, death, or last follow up, whichever occurred first. All analysis was performed in R version 4.4.0 and two-sided p-values less than 0.05 were considered statistically significant.

### Genomics

#### Sample Preparation and Sequencing

DNA samples were extracted from each tumoroid using the Qiagen AllPrep DNA/RNA Kit per kit protocol. Twelve tumoroids derived from eleven rectal cancer patients underwent whole exome sequencing (WES) alongside patient-matched germline (blood) samples. For the patient with four tumoroids derived at different time points, only the primary tumor-derived tumoroid was included in this analysis to mitigate over-representation of one patient. The tumoroids were sequenced at an average depth of 150×, and each patient’s corresponding blood sample was sequenced at an average depth of 70× at the Memorial Sloan Kettering Cancer Center (MSKCC). All raw sequencing data underwent standardized processing using the TEMPO pipeline [(https://github.com/mskcc/tempo)].

The four tumoroids representing the disease progression of a single patient were then subjected to WES at the John P. Hussman Institute for Human Genomics (HIHG) at the University of Miami, achieving an average depth of 200×. To control for potential batch effects, the primary tumor sample was re-sequenced at HIHG under the same conditions. Raw FASTQ files for these four samples were processed using the nf-core Sarek pipeline[14] (version 3.4.4) on Nextflow[15] (version 24.04.4) with default parameters and the “--wes” option.

#### Variant Calling and Filtering

For all tumoroids, somatic variants were called using Mutect2 and Strelka2 integrated in Sarek. Variants annotated as “PASS” by both callers were retained for downstream analysis. To minimize artifactual signals, recurrent technical artifacts were filtered using a combination of the GATK Panel of Normals (PoN; MuTect2.PON.5210.vcf.tar) and a custom PoN. Only variants surviving these filters were considered for subsequent steps.

#### Copy Number Analysis

FACETS[16] was applied to infer tumor purity and gene-level copy number (CN) alterations. This was performed using the facetsSuite package (version 2.0.8; https://github.com/mskcc/facets-suite) in R[17] (version 4.3.2), providing tumor purity estimates and CN segmentation profiles for downstream integrations.

#### Significantly Mutated Genes and Mutational Signatures Analysis

To identify significantly mutated genes across twelve tumoroids, we ran MutSig2CV[18] (https://github.com/getzlab/MutSig2CV) in MATLAB[19] (R2024a), based on somatic SNPs and indels using the human reference genome hg19 as the reference. For the patient with four tumoroids derived at different time points, only the primary tumor-derived tumoroid was included in this analysis to mitigate over-representation of one patient. Significant genes (Fisher’s combined p-value < 0.01) containing non-silent mutations with “high” or “moderate” impact as annotated by SnpEff.[20] based on the Ensembl[21] release 112 database (https://may2024.archive.ensembl.org) were selected for oncoprint visualization. The oncoprint was generated by the ComplexHeatmap[22] package (version 2.18.0; https://github.com/jokergoo/ComplexHeatmap) in R.

We further explored single base substitution (SBS) mutational signatures using tempoSig (version 0.5.4; https://github.com/mskcc/tempoSig). SBS signatures defined by COSMIC[23] (version 3; https://cancer.sanger.ac.uk/signatures/sbs/) were decomposed for each sample, generating a matrix of estimated signature fractions and corresponding p-values derived from 1,000 permutation tests. The relative contributions of each SBS signature were subsequently modeled via linear regression against intrinsic radiosensitivity values.

#### Phylogenetic Analysis

To reconstruct clonal architectures and infer phylogenetic relationships among the four tumoroids representing the disease progression of a single patient, we aggregated the union of non-silent somatic SNVs identified across all samples. Reference and variant allele counts were obtained using bam-readcount (version 1.0.1; https://github.com/genome/bam-readcount) with parameters -q 10 -b 30 -w 0. The resulting read counts were processed using the bamreadcountr R scripts (https://github.com/sahilseth/bamreadcountr). Cancer cell fraction (CCF) estimates for each variant were inferred using ABSOLUTE[24], facilitated by the DoAbsolute wrapper (Release 2.2; https://github.com/ShixiangWang/DoAbsolute) in R. These estimates were informed by tumor purity and gene-level CN data from FACETS.

PhylogicNDT[25] (version 1.0; https://github.com/getzlab/PhylogicNDT) was then executed under Python 2.7[26] to cluster mutations into clonal and subclonal populations, construct phylogenetic trees, and quantify the clonal architecture. Default parameters were used, except for setting the number of iterations (-ni) to 1,000. The most frequently observed phylogenetic tree was selected, and the posterior CCF distributions from PhylogicNDT were used to finalize clonal/subclonal proportions within each sample.

## Results

Fifteen tumoroids and three normal rectal tissue-derived organoids were studied, representing eleven patients. Three patients were female, and eight were male. Median age at tumoroid derivation was 45 years, ranging from 29 to 74 years. Stage I, III, and IV disease was represented. One patient was MSI-H. Patient demographics and sample data are shown in **Figure 1A**.

**Figure 1:**
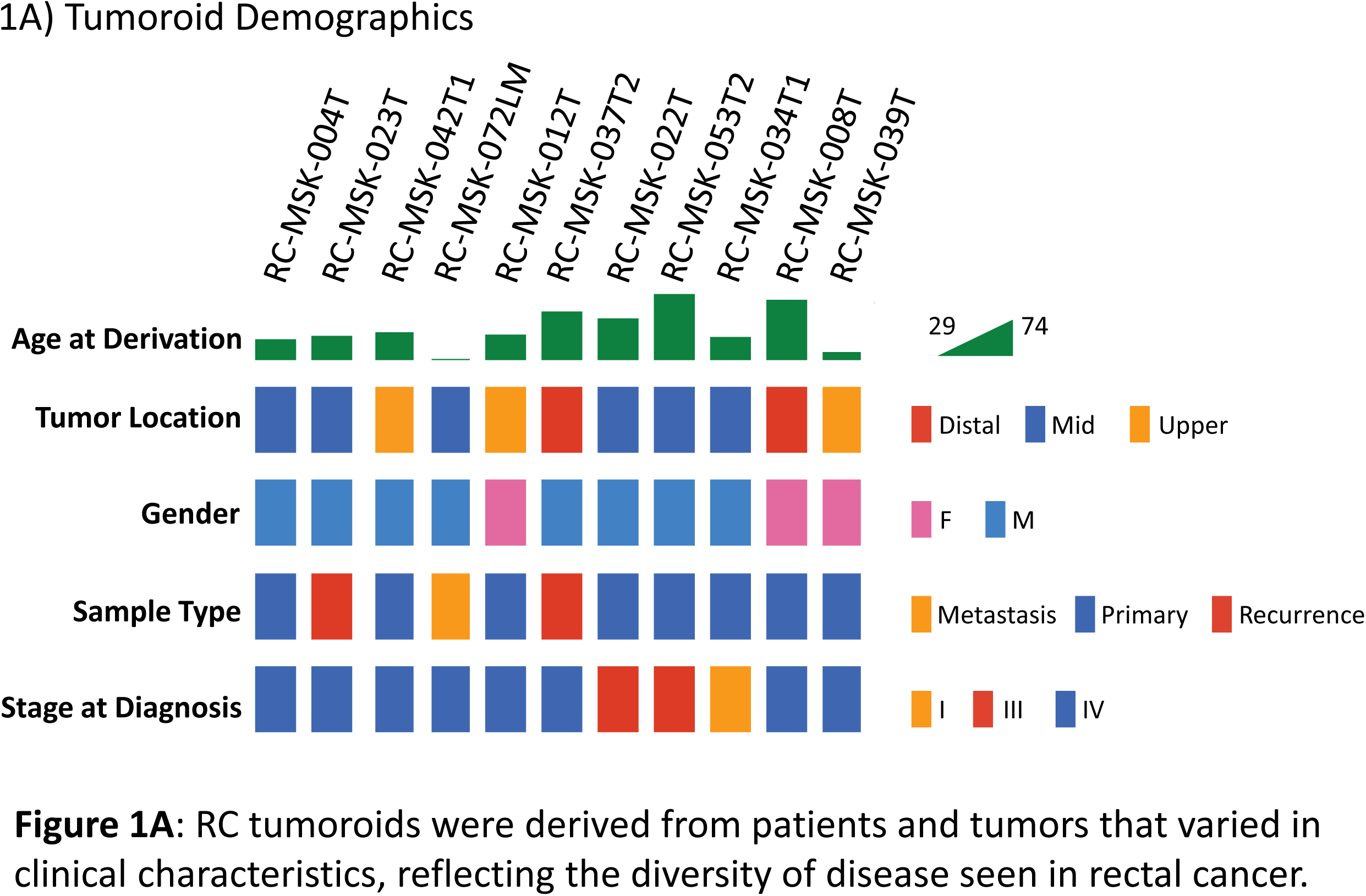
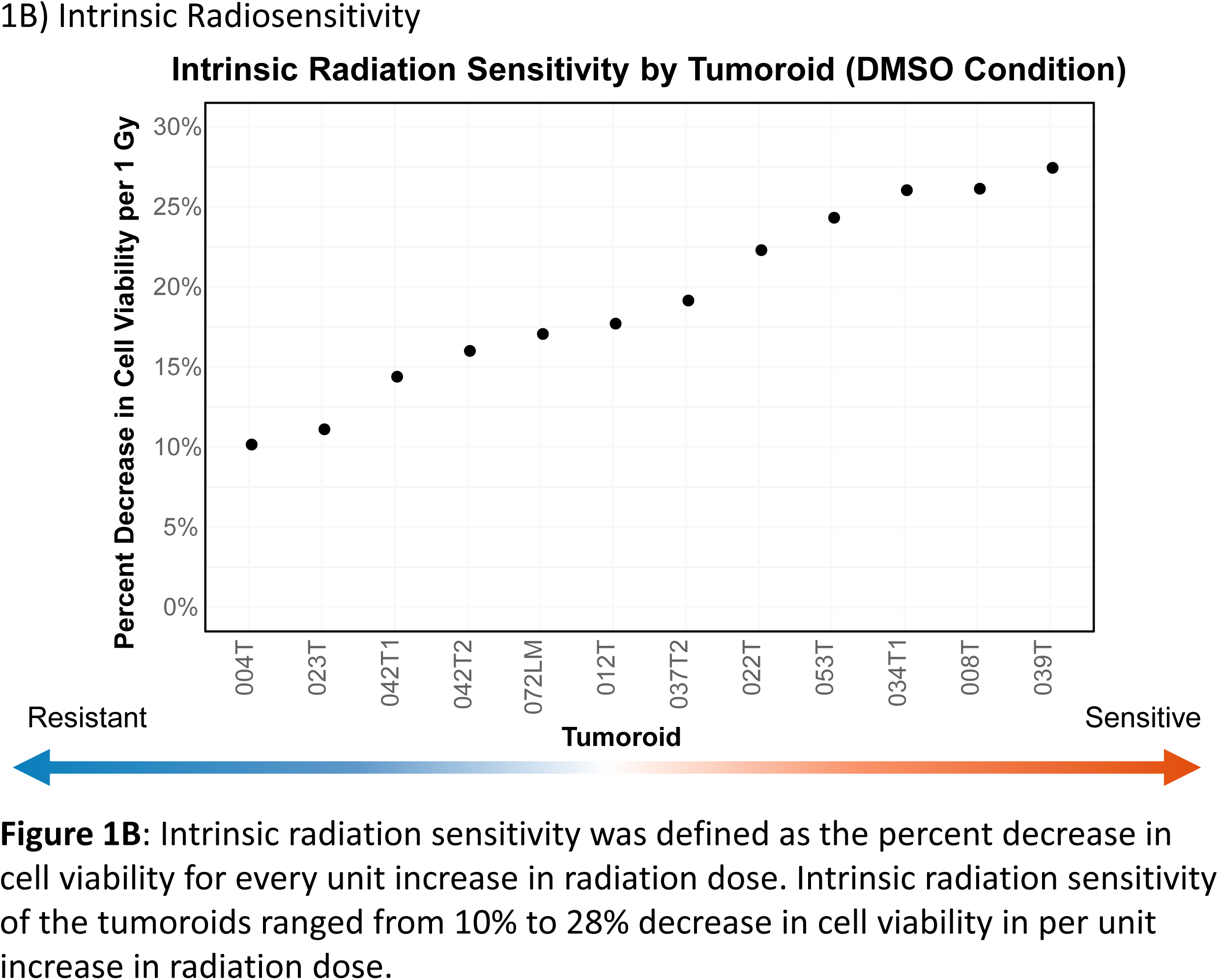
RC tumoroids reflecting a diversity of disease also vary in intrinsic radiation sensitivity.

### Radiation Sensitivity and Sensitization

Tumoroids demonstrated intrinsic differences in sensitivity to radiation when irradiated in the absence of drug treatment (DMSO only). The most radiosensitive tumoroid was RC-MSK-039T with a 28% decrease in cell viability for every unit increase in radiation dose. Conversely, the most radioresistant tumoroid was RC-MSK-004T with only a 10% decrease in cell viability for every unit increase in radiation dose. Tumoroids derived from the same patient at different tumor sites also varied in their radiosensitivity relative to each other: tumoroids RC-MSK-042T1 and RC-MSK-42T2 were biopsied at the same time from heterogeneous areas of the same tumor and demonstrated a small difference in decreasing cell viability (14.5% vs 16.2%, respectively). **Figure 1B** displays the intrinsic radiosensitivity for each tumoroid (listed in Supplementary Table 3). Thus, tumoroids displayed heterogeneity in their intrinsic radiosensitivity, paralleling the clinical responses seen in rectal cancer.

Tumoroids were roughly characterized as sensitive or resistant along the median value of intrinsic radiation sensitivity. Patients whose tumoroids were characterized as radiation-resistant (n=5) had a four-year overall survival (OS) of 20% (95% CI: 3.5%, 100%) and all had a progression event, with a median progression-free survival (PFS) of 9.3 months (95% CI: 4.3, -). Conversely, patients who had radiation-sensitive tumoroids (n=6) had a four-year OS of 50% (95% CI: 22%, 100%) and a median PFS of 16 months (95% CI: 7.8, -). Survival curves can be seen in Supplementary Figure 2.

When treated with DDRis, the radiation sensitivities of the tumoroids were altered from the baseline DMSO condition. For example, **Figure 2A** demonstrates the response of RC-MSK-012T to drug treatment in a log linear cell viability plot and illustrates that DNA-PKi and ATMi increased cell kill (or conversely, decreased cell viability) as radiation dose increased. Linear cell viability plots for every tumoroid are shown in Supplementary Figure 3. Immunohistochemistry and Jess Automated Western Blot confirmed inhibition of downstream targets with drug treatment and subsequent radiation (**Figure 2B-2D**, Supplementary Figure 4).

**Figure 2:**
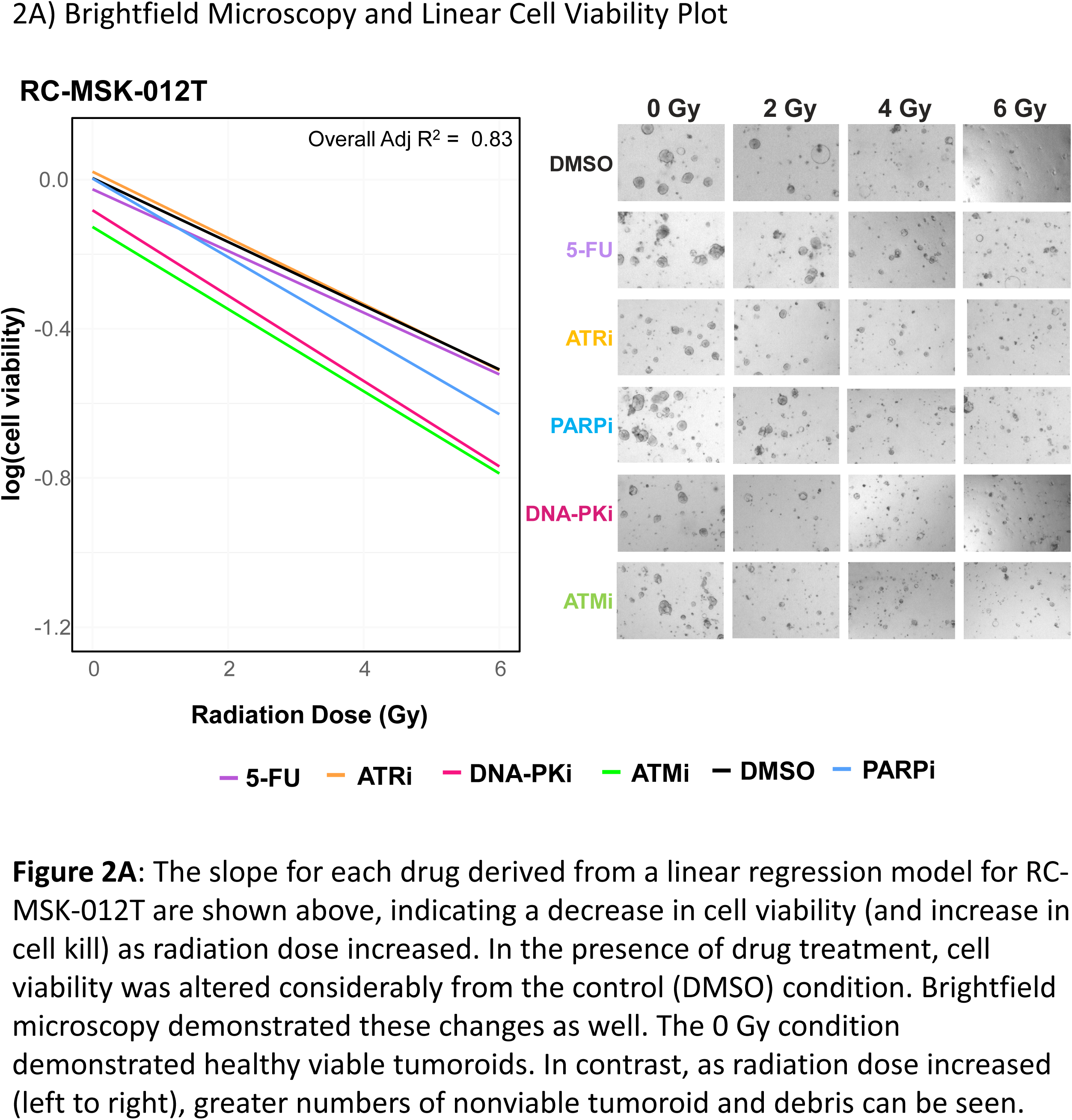
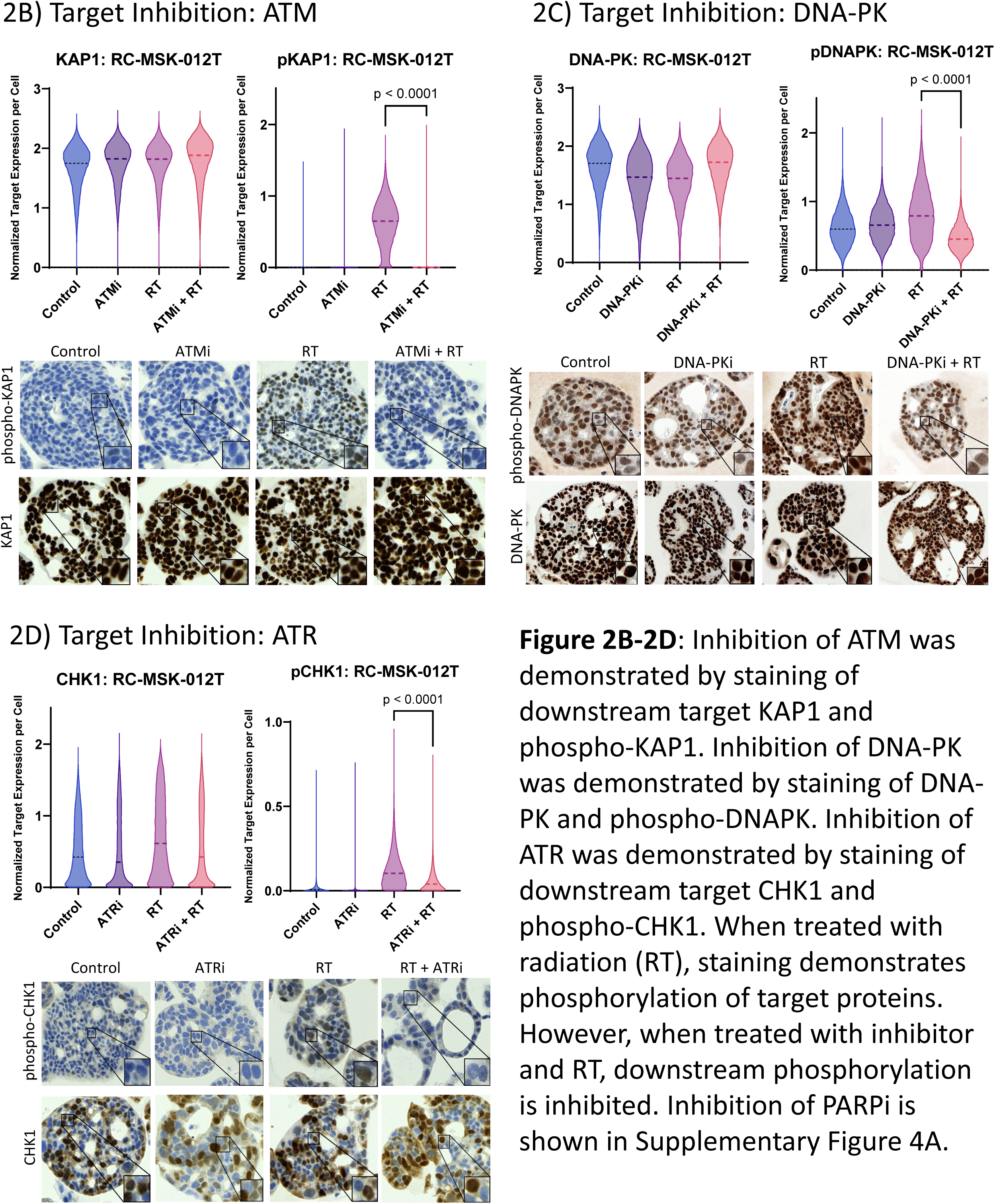
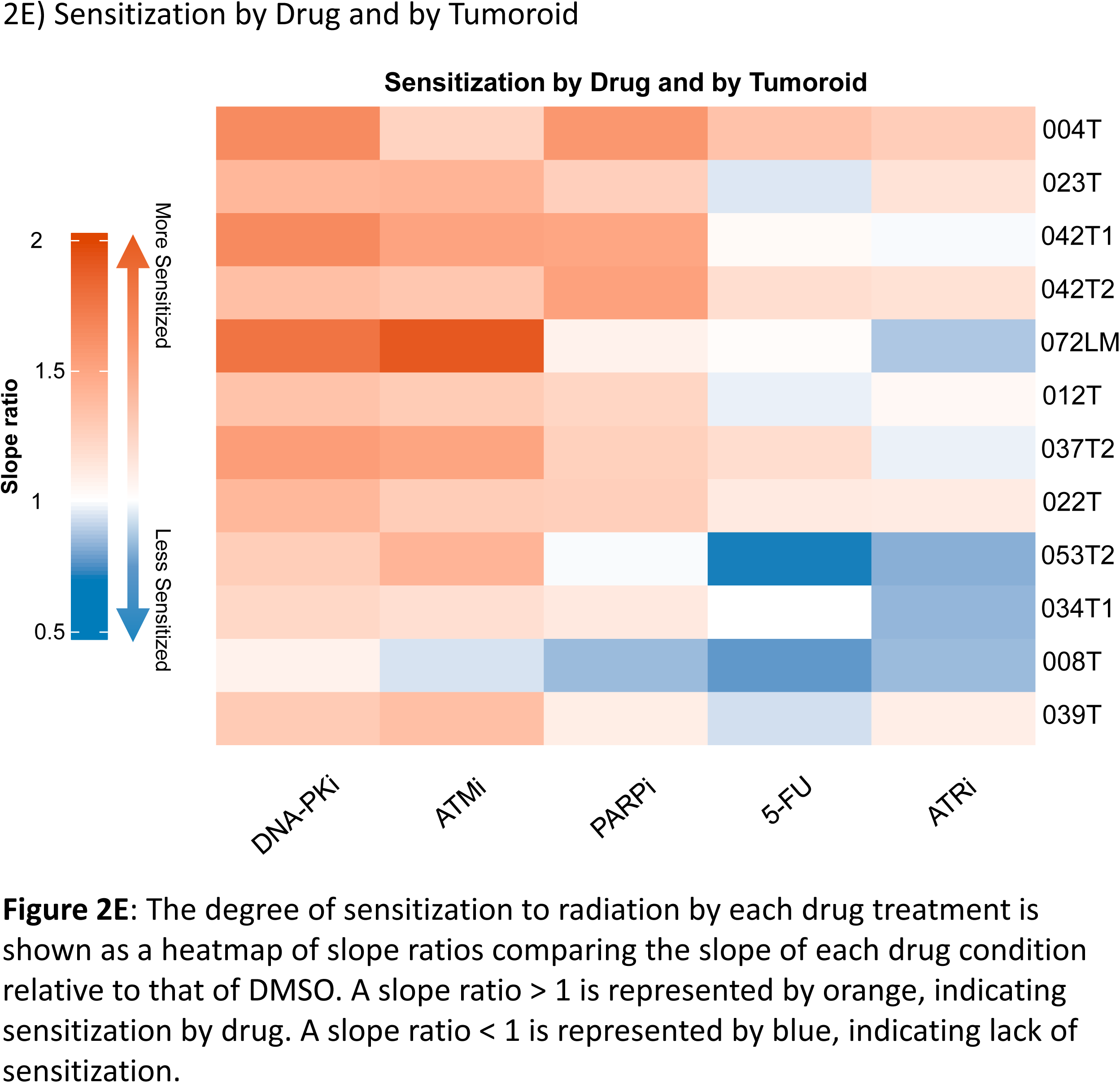
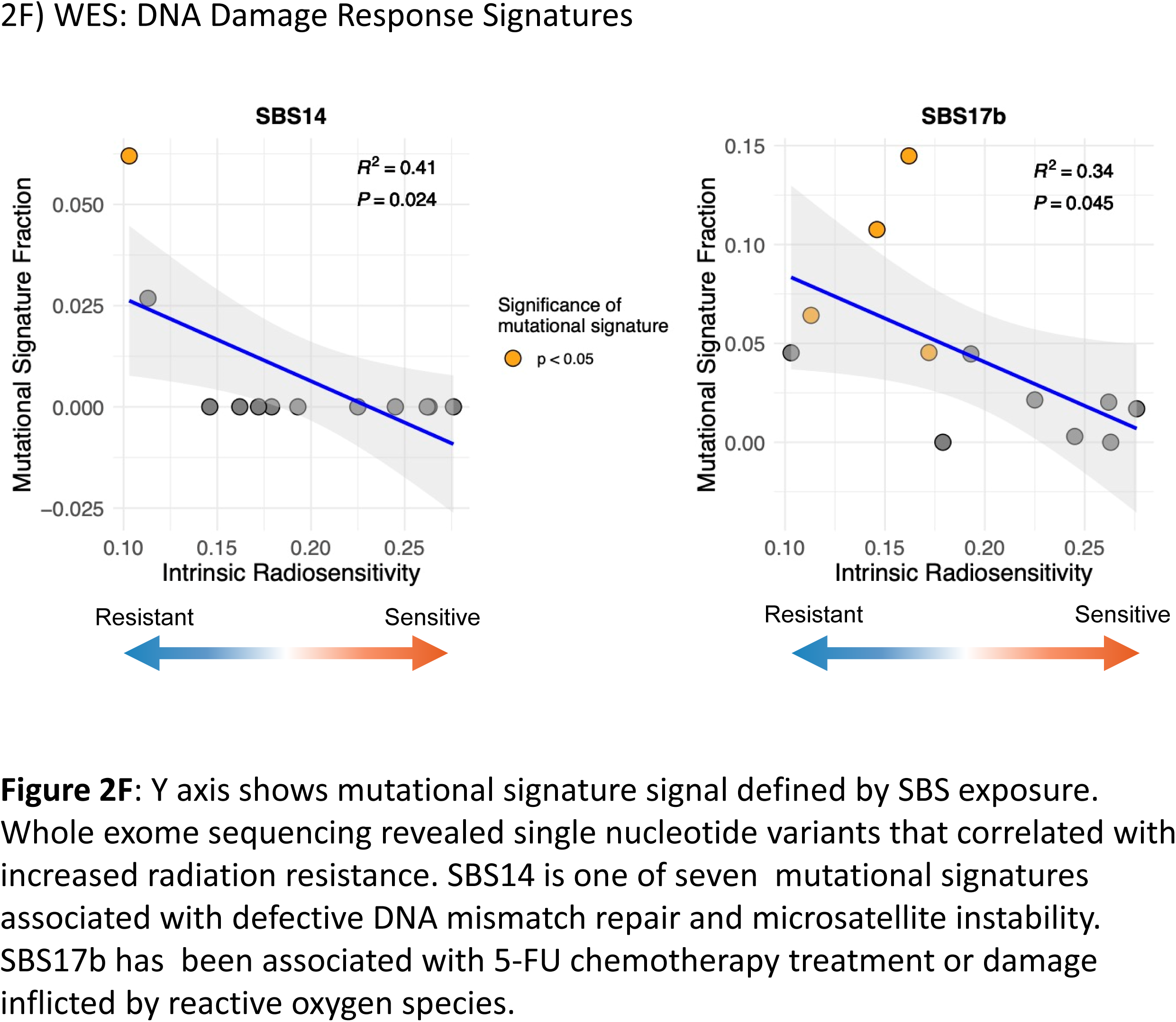
RC tumoroids can be sensitized using DNA damage repair inhibitors.

Overall, between tumoroids, the degree of sensitization to radiation by drug treatment displayed substantial heterogeneity as well. **Figure 2E** displays a heatmap of slope ratios, demonstrating that the degree of sensitization varies by both tumoroid and by drug choice (tabulated in Supplementary Table 4). Relative to DMSO, the most potent radiosensitizer was DNA-PKi in 7 tumoroids, ATMi in 4 tumoroids, and PARPi in 1 tumoroid. ATRi and 5-FU were the least effective radiosensitizers. Intrinsically radiation-resistant tumoroids showed a trend toward greater sensitization.

### Mutational Landscapes of Patient-Derived Tumoroids

WES of twelve tumoroids from eleven patients uncovered mutation profiles characteristic of colorectal cancer (Supplementary Figure 5). Among the 44 significantly mutated genes (MutSig2CV p-value < 0.01), APC, TP53, and KRAS were most frequently affected, with non-silent mutations in APC present in 92% of patients, followed by TP53 (67%) and KRAS (58%). Despite the focus on radiation sensitivity, no significantly altered somatic mutations were detected in the DNA damage repair pathway gene except for TP53.

We next examined associations between SBS mutational signatures and intrinsic radiosensitivity. Each SBS signature fraction – derived from a 67-signature decomposition – was modeled against the intrinsic radiation sensitivity (i.e., 1 – β metric) using a linear regression. Two signatures, SBS14 and SBS17b, showed significant negative correlations with radiation sensitivity (p-value < 0.05). Notably, SBS17b was present at a significant fraction (tempoSig p-value < 0.05) in more tumoroids than SBS14, albeit based on a limited sample size (**Figure 2F**). Thus, SBS17b and SBS14 could be correlated significantly with increased radiation resistance. Of note, SBS14 is one of seven mutational signatures associated with defective DNA mismatch repair and microsatellite instability.[27] SBS17b has been associated with 5-FU chemotherapy treatment or damage inflicted by reactive oxygen species.[28]

### Comparing Tumoroids in Series

Patient RC-MSK-012 had four tumoroids derived over their treatment course, representing different disease timepoints. Two tumoroids were derived after TNT at the time of index surgery from the primary rectal tumor (RC-MSK-012T) and a splenic metastasis (RC-MSK-012SP). Four months later, after additional chemotherapy, the patient’s disease progressed with rectal stump and vaginal wall recurrences. These recurrences were also derived into tumoroids (RC-MSK-012PR and RC-MSK-012VR, respectively).

Intrinsic sensitivity to radiation (DMSO condition) was similar in all four tumoroids, with slightly greater radioresistance in post-progression tumoroids. However, sensitization by drug treatment was more pronounced in pre-progression tumoroids as compared to post-progression tumoroids derived from recurrences (**Figure 3A**). A more granular look at RC-MSK-012T (pre-progression) and RC-MSK-012PR (post-progression) demonstrates steeper slopes with drug treatment in the pre-progression tumoroid than the post-progression tumoroid (**Figure 3B**). These findings reflect a loss of sensitization with drug treatment in the post-progression tumoroid. Assessing the effects on downstream targets through immunohistochemistry and Jess Automated Western Blot confirmed a relative loss of target inhibition in the post-progression tumoroid compared to the pre-progression tumoroid (**Figure 3C**, Supplementary Figure 4). Thus, post-progression tumoroids demonstrated greater radioresistance and diminished DDRi-induced radiosensitization.

**Figure 3:**
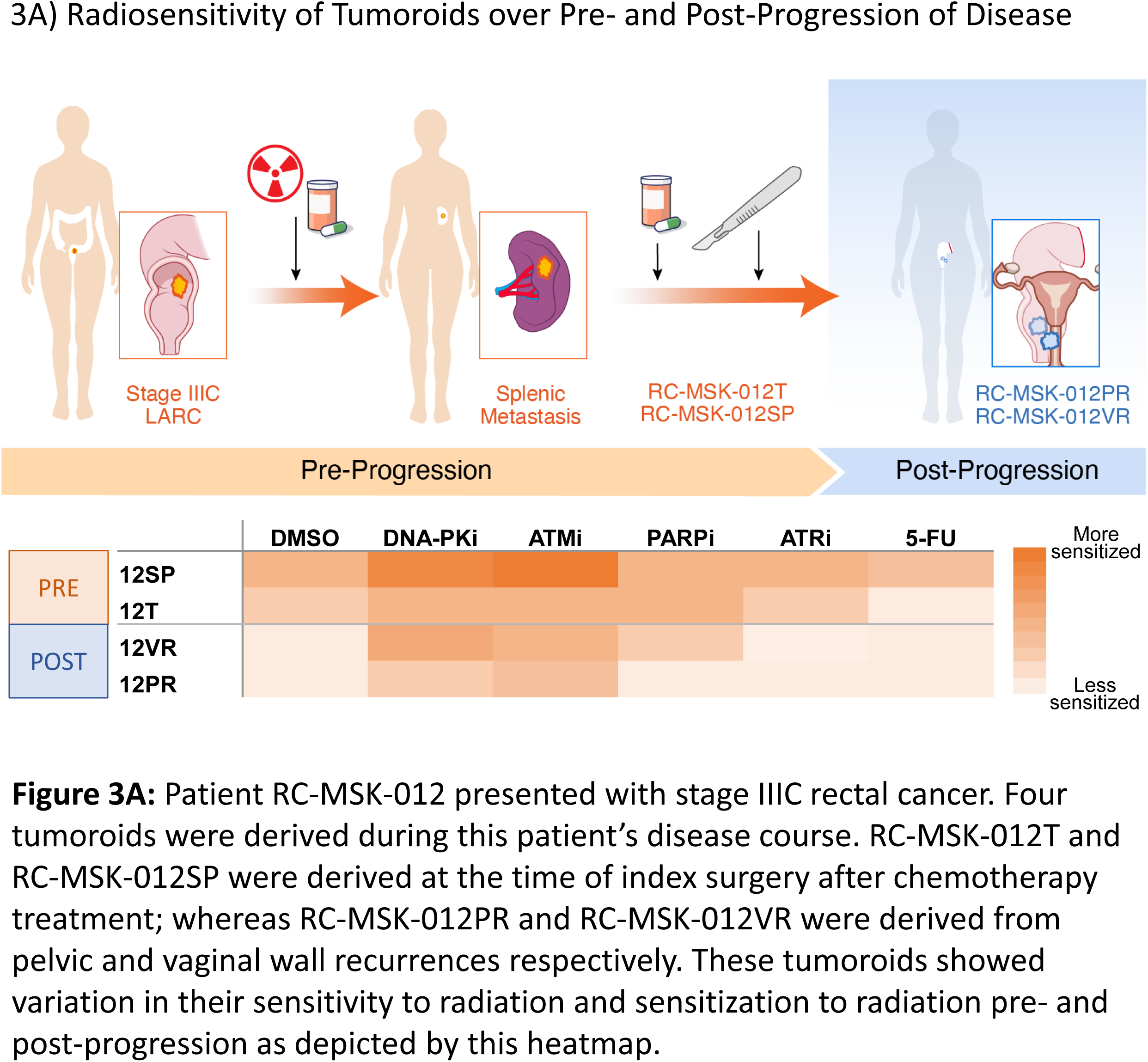
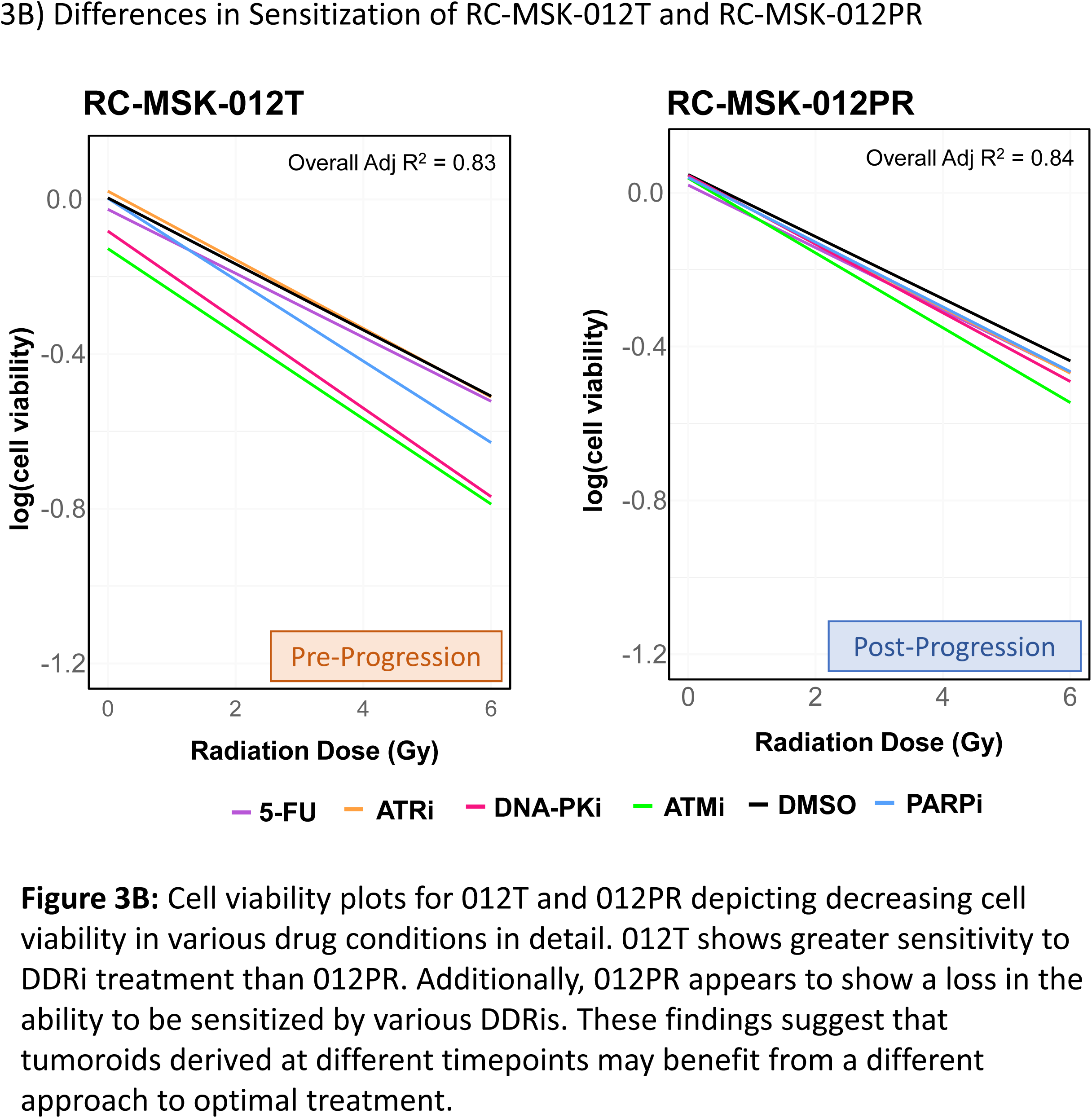
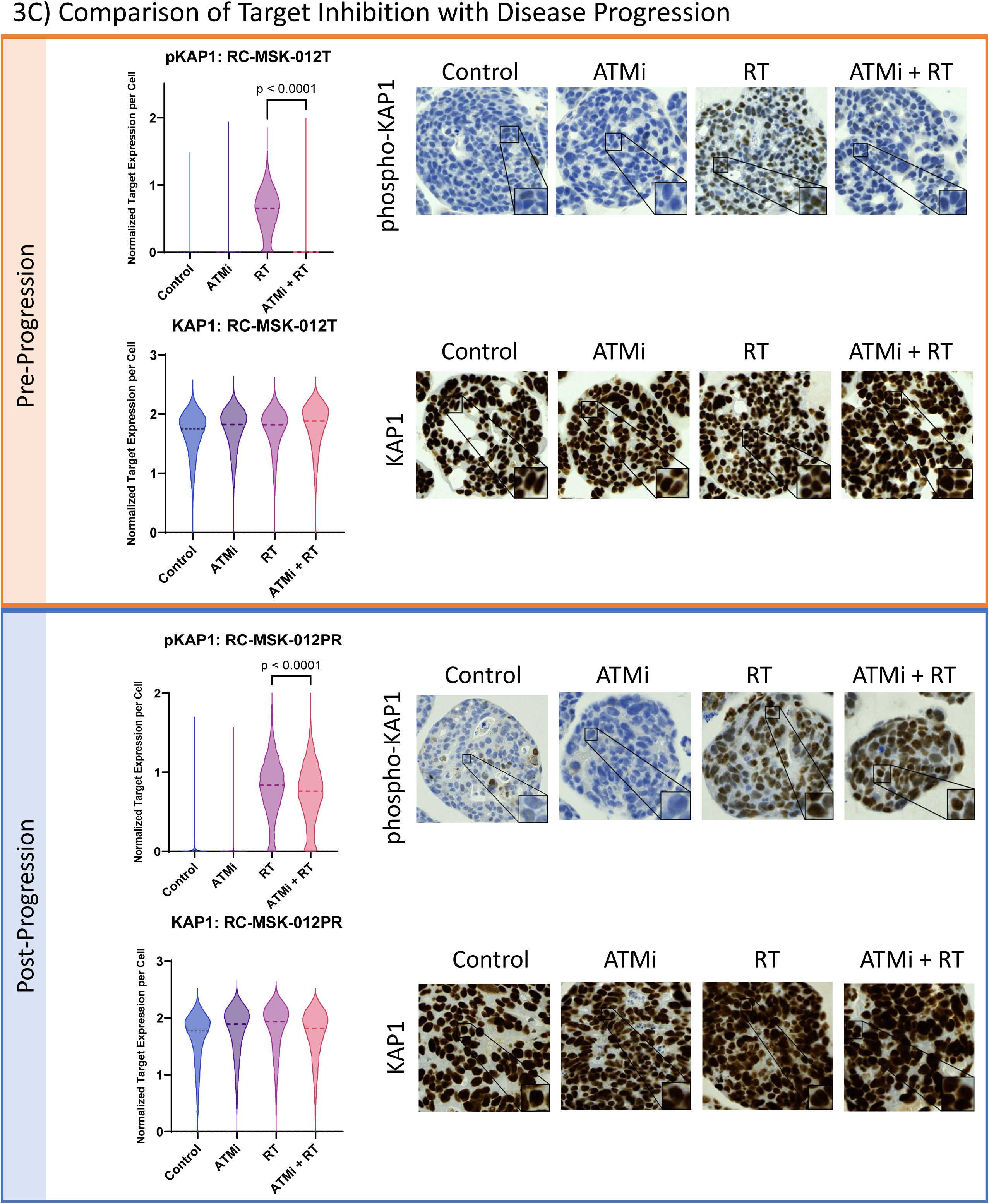
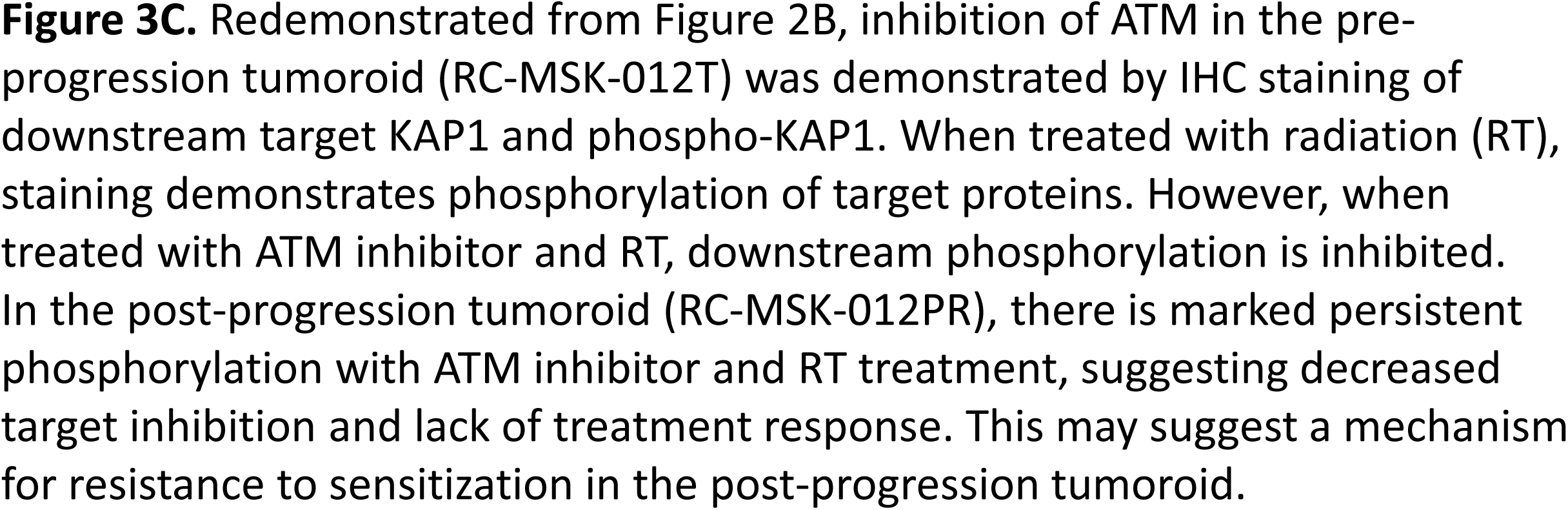
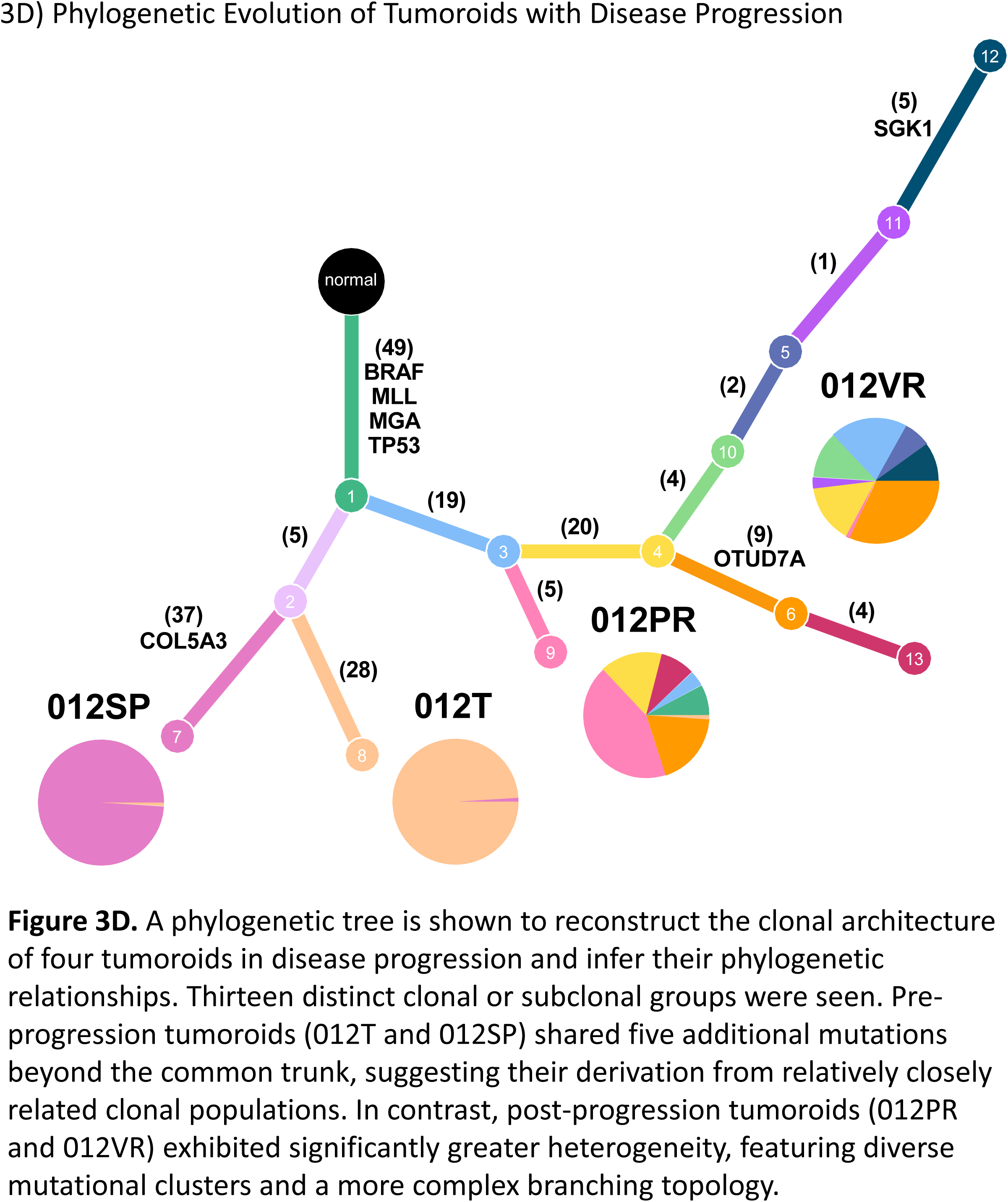
The Sensitizing Effects of Radiosensitizers Change with Disease Progression.

### Phylogenetic Analysis of Tumoroids over Disease Progression

Given that the most patients with rectal cancer not cured by TNT die from progressive and metastatic disease, we elected to further investigate the unique set of tumoroids derived from a primary tumor and matched recurrent and metastatic samples.[29, 30] WES analysis of the primary tumor-derived tumoroid (012T) and three metastasis- or recurrence-derived tumoroids (012SP, 012PR, and 012VR) revealed an evolving mutational landscape (**Figure 3D**). In total, 188 non-silent SNVs and indels were identified across these four samples, and PhylogicNDT clustering resulted in thirteen distinct clonal or subclonal groups.

Constructing a phylogenetic tree from these clusters demonstrated that only 49 (26%) of the identified mutations were present in all four tumoroids, indicating substantial genetic divergence during disease progression. Among the lineages, 012T and 012SP shared five additional mutations beyond the common trunk, suggesting derivation from closely-related clonal populations. In contrast, 012PR and 012VR exhibited greater heterogeneity, featuring diverse mutational clusters and a more complex branching topology. Examining the evolution of tumors with disease progression and their potential mechanisms for resistance provides a path toward the adaptation of treatment to progressive, recurrent, and likely treatment-resistant, disease.

### Comparing Tumoroids to Normal Tissue-Derived Organoids

Given a complete lack of models for studying normal/tumor biology and expansion of the therapeutic index in rectal cancer and gastrointestinal tumors, we sought to amplify our tumoroid system to accomplish three goals: a) establish a robust normal/tumor rectal cancer platform for precision therapy; b) expand this platform to identify and exploit the therapeutic window on a patient-specific level; and c) provide proof-of-principle to identify radiosensitizers that could preferentially be used to avoid toxicity in normal tissues in future Phase I clinical trials.

To this end, we examined tumor-selective sensitization (**Figure 4A**), assessed by comparing the cell viability of three tumoroids to their matched normal organoids in each treatment condition. For example, the decrease in cell viability for RC-MSK-012T (tumoroid) in the PARPi condition was greater than the decrease in cell viability for RC-MSK-012N (organoid) in the PARPi condition, as demonstrated by the slopes of each curve (**Figure 4B**). These data show greater sensitization by drug treatment in the tumoroid than the organoid for all drug conditions.

**Figure 4:**
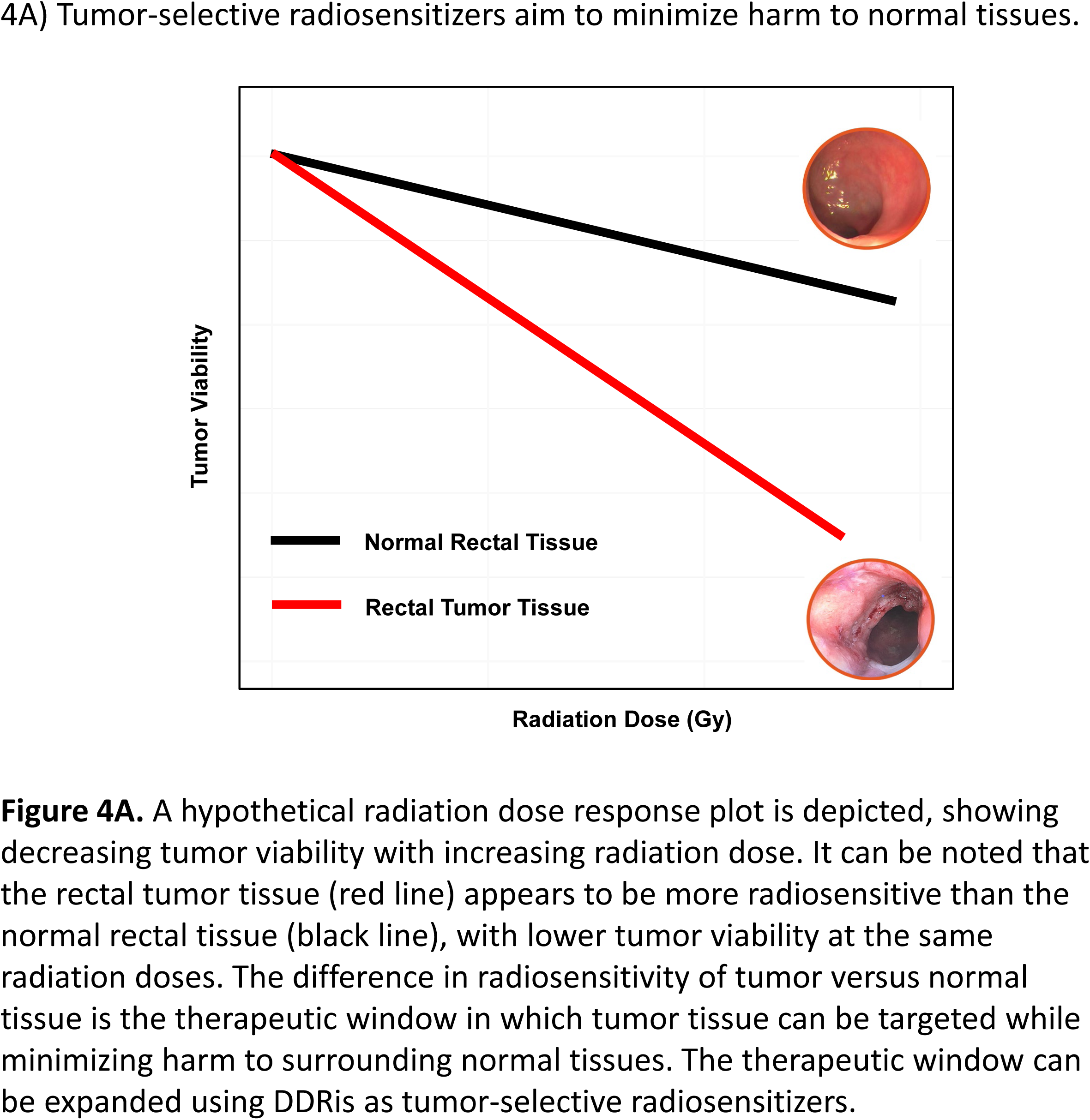
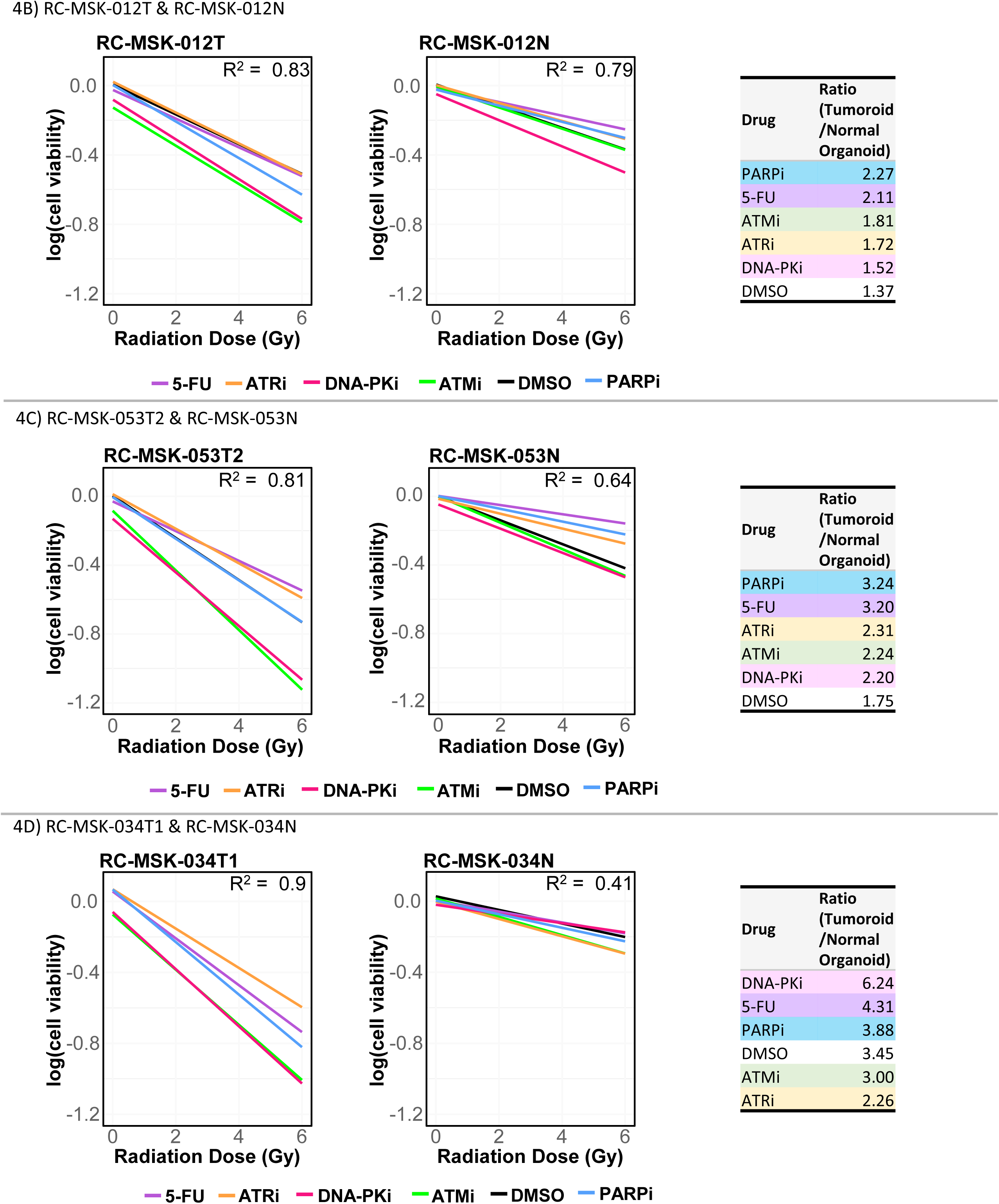
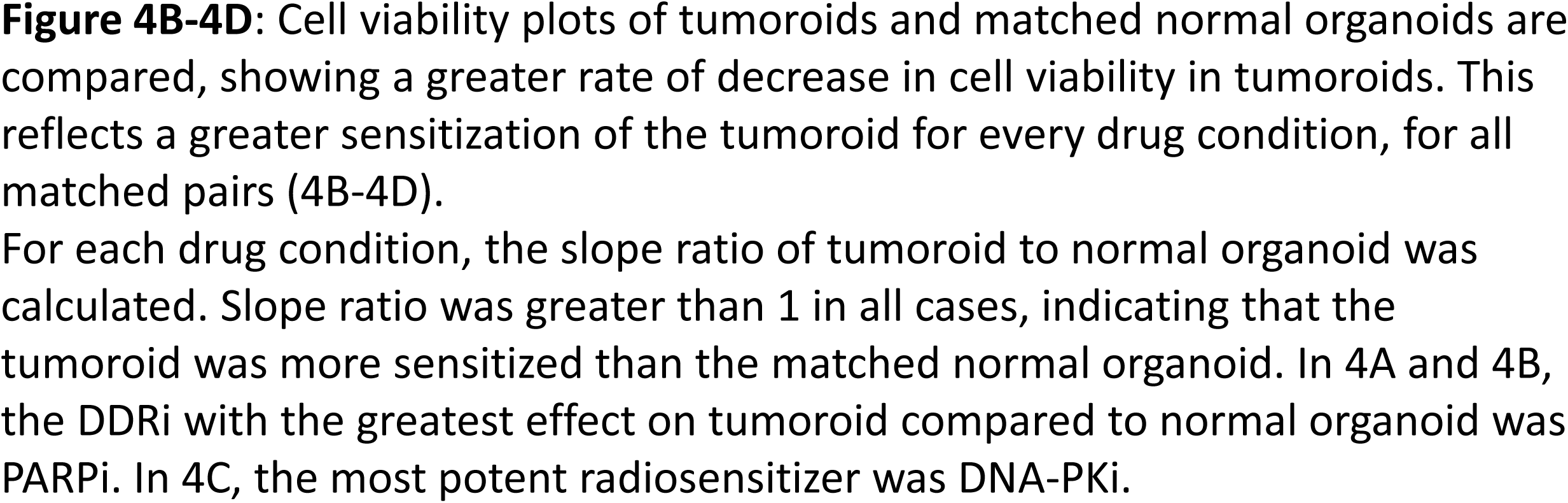
RC tumoroids showed greater sensitization to radiation than their corresponding matched normal organoids.

Consistently, the slopes of the tumoroid plots showed a greater rate of decrease in cell viability compared to normal organoid, reflecting greater sensitization by drug treatment for all matched pairs (**Figure 4B-D**). Next, the slope of each tumoroid treatment condition was compared to the corresponding organoid treatment condition to yield a tumoroid-to-normal organoid ratio. In all cases, the slope ratio was greater than 1, indicating that the tumoroid was more sensitized than the matched normal organoid. For RC-MSK-012T and RC-MSK-053T2, the DDRi with the greatest effect on tumoroid relative to normal organoid was PARPi. For RC-MSK-034T1, the most potent radiosensitizer was DNA-PKi.

## Discussion

Chemoradiation is a cornerstone in the treatment of rectal cancer; however, the efficacy of radiation therapy is often limited by differing tumor responses among patients. In this study, a rectal cancer tumoroid platform was used to recapitulate this recognized clinical variation in an *ex vivo* model. To our knowledge, this is the first study to quantitatively characterize intrinsic radiosensitivity across a spectrum of tumoroids ranging from radiosensitive to radioresistant. Notably, the utility of this model even extends to describing heterogeneity within a single tumor (RC-MSK-042), as well as enabling the longitudinal study of tumoroids at distinct disease timepoints (RC-MSK-012). In addition to intrinsic radiosensitivity, the tumoroid platform was also used to study the effects of various DNA damage repair inhibitors (DDRis) as radiosensitizers. The DDRi with the most potent sensitizing effect appeared to vary by tumoroid, again reflecting the heterogeneity seen clinically in rectal cancers. Finally, we establish for the first time a normal/tumor platform to better investigate and exploit the therapeutic window for future studies, allowing for the selection of potent radiosensitizers that spare toxicity to normal tissues. These data suggest that the selection of DDRis could be more precisely tailored to match an individual patient’s tumor and physiology. Moreover, our tumoroid platform offers a method by which to efficiently assess the efficacy of these drugs *ex vivo* for individual patients.

A key advantage of our tumoroid biorepository platform is the unique capacity to store and analyze patient -derived tumoroids over time, capturing changes that arise with treatment or disease progression. In this study, the progression of disease and its effect on intrinsic radiation sensitivity and sensitization was investigated using four tumoroids in series from a single patient (**Figure 3**). Two tumoroids represented pre-progression disease (RC-MSK-012T and RC-MSK-012SP) and two tumoroids represented post-progression disease (RC-MSK-012PR and RC-MSK-012VR). While sensitivity to radiation in the absence of drug treatment (DMSO only) was similar in all four tumoroids, the post-progression tumoroids reflected a relative loss in sensitization by certain DDRis. The loss of target inhibition was similarly noted on immunohistochemistry. Phylogenetic studies of these four tumoroids demonstrated substantial genetic divergence during disease progression and markedly increased heterogeneity in post-progression tumoroids. While this analysis focused on evolutionary relationships, ongoing efforts aim to highlight specific genes of interest that may drive these phylogenetic patterns or influence therapeutic vulnerabilities. Characterization of tumoroids through disease progression can illuminate both mechanisms of resistance and opportunities to refine subsequent therapies for patients who do progress.

Through the investigation of DDRis in rectal cancer, we aim to optimize radiation therapy and TNT to improve cCR rates and increase the number of patients who can benefit from WW surveillance. Though multiple TNT regimens are available with demonstrated effectiveness for many patients, the radiation component of these therapies continues to have adverse effects. Further, specific groups of patients may derive additional benefit from the application of the tumoroid model or adjunctive DDRi treatment, such as those with clinically radioresistant tumors, those who are unable to tolerate radiation therapy due to adverse effects, and those who progress after undergoing recommended therapy and have limited current options for salvage. For example, this tumoroid model can identify patients who may have tumors that are intrinsically very resistant to radiation therapy. These patients may benefit from the omission of radiation therapy. Understanding the limitations of radiation treatment in certain resistant tumors may allow clinicians to one day redirect these patients to chemotherapy-escalation approaches early in the treatment course and avoid the adverse treatment effects of radiation therapy. Limiting radiation therapy has been studied in the PROSPECT trial (NCT01515787) and is ongoing in the NORAD01-GRECCAR16 trial (NCT03875781); however, it has not been studied in the context of WW and prospective clinical trials would be needed to consider the selective omission of chemoradiation in LARC patients.[31, 32] Additionally, our tumoroid platform could also benefit patients suffering from the adverse effects of radiation therapy. Many of these patients are unable to complete the full course of radiation therapy. For these patients, a DDRi that sensitizes their tumor to radiation may allow for the use of lower doses of radiation, sparing their normal tissues from prohibitive damage without fully compromising tumor cell apoptosis. Thus, more patients may be able to complete TNT in its entirety, offering the best chance for cCR and reduced operative morbidity.

Despite these promising insights, this study’s small cohort size limited the scope of our conclusions regarding radiosensitivity, DDRi efficacy, and genomic correlates. Fifteen tumoroids from eleven patients were analyzed, with only three tumoroid-normal organoid pairs and only one patient with serial samples. Importantly, the tumoroid cohort should be expanded to include additional stages of disease and patients of different ethnic backgrounds (specifically African American patients) more comprehensively. Moreover, no single DDRi emerged as a universally superior treatment option; however, the variability in response underscores the utility of an individualized testing platform for refining treatment choices. Additionally, it lends credence to the flexibility of the tumoroid platform to investigate patients individually and at different disease time points. Future directions include additional studies of tumoroid-normal organoid pairs and tumoroids in series to describe these relationships more completely. Based on this work, we have subsequently established normal/tumor pairs in an additional 329 samples as candidates for future experiments (parallel paper in progress, data not shown). Further transcriptomic studies are also ongoing and could be utilized to point to biomarkers that help predict radiation sensitivity or molecular targets that can be used to sensitize tumoroids.

In conclusion, this is the first ex vivo study to quantify intrinsic radiation sensitivity and DDRi sensitization in a patient-specific tumoroid platform. To our knowledge, this is the first study to evaluate radiation sensitivity in this way. Because the heterogeneity seen in radiosensitivity is not fully explained by mutational differences, this platform underscores the utility of a patient-matched tumoroid-organoid model as a platform to quantify and predict personalized responses to radiation and DDR inhibitors. Moreover, by comparing tumoroids from a single patient across different disease stages, our model illustrates how tumors evolve under treatment pressure and become more radiation resistant, offering a potential framework to inform future precision radiotherapy strategies. Finally, this model demonstrates that radiosensitizer efficacy is tumor-selective, with important clinical implications for minimizing harm to normal tissues. In the modern treatment paradigm of rectal cancer with TNT and selective WW, our platform highlights opportunities for and a method by which to refine clinical treatment for patients with radioresistant tumors, minimize normal tissue injury, and adapt therapy over disease course. Further, we offer a robust model that accurately recapitulates tumor biology and heterogeneity, serving as a platform that can potentially be expanded to various malignancies.

## Data Availability

All data produced in the present study are available upon reasonable request to the authors.

## Funding

This work was supported in part by a National Institutes of Health/National Cancer Institute, NIH/NCI) Memorial Sloan Kettering Cancer Center (MSK) support grant (P30CA008748). Dr. Wini Zambare was supported by NCI Surgical Oncology T32 research training grant (T32CA9501-34). Dr. Smith is supported by an NIH/NCI grant (R37CA248289), the Department of Surgery and Colorectal Service at MSK (J. Drebin/J. Garcia-Aguilar), the Wasserman Colorectal Research Fund, and generous donations by Corinne Berezuk, Michael Stieber, and Patrick A. Gerschel (Dr. Paty). Dr. Romesser is supported by an NIH/NCI grant (K08CA255574), EMD Serono research grant, and an MSK Imaging and Radiation Sciences Program (IMRAS) grant, and a Geoffrey Beene Cancer Research Center grant. Finally, this research was also supported by the Sylvester Comprehensive Cancer Center at the University of Miami Miller School of Medicine which receives funding from the National Cancer Institute award P30CA240139 (Dr. Chen).

## Conflicts of Interest

P.B.R. received research funding (2019) and served as a consultant for EMD Serono (2018-2024), receives research funding from XRAD Therapeutics (2022-present), is a consultant for Faeth Therapeutics (2022-present), is a consultant for Natera (2022-present), and is a volunteer on the advisory board for the HPV Alliance and Anal Cancer Foundation non-profit organizations.

J.J.S. received travel support for fellow education from Intuitive Surgical (August 2015). He also served as a clinical advisor for Guardant Health (March 2019) and as a clinical advisor for Foundation Medicine (April 2022). He served as a consultant and speaker for Johnson and Johnson (May 2022). He served as a clinical advisor and consultant for GlaxoSmithKline (2023-24).

## Acknowledgements

The authors would like to thank the Colorectal Disease Management Team at Memorial Sloan Kettering for their support.

## Supplementary Figures

**Supplementary Figure 1:**
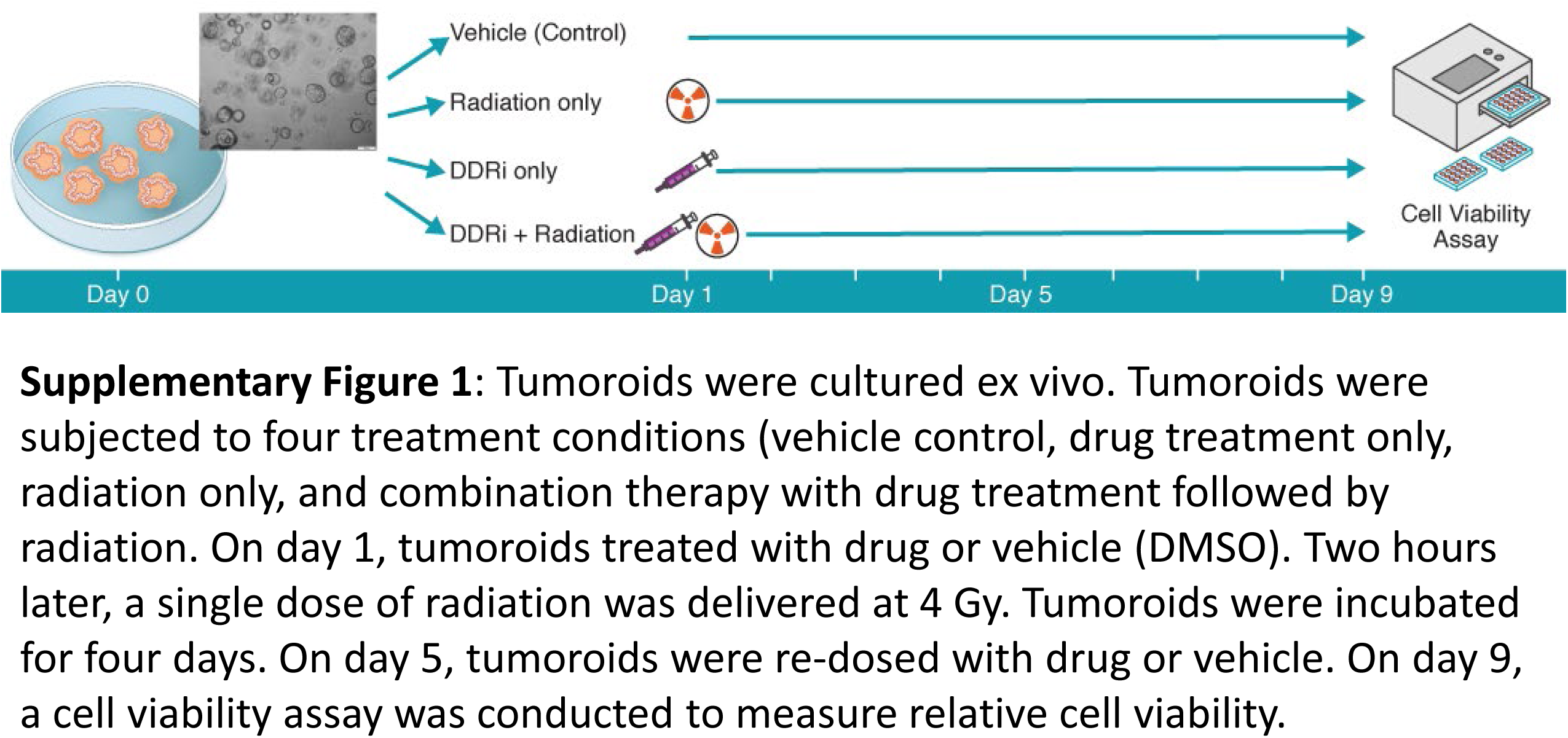
Schematic of Methods.

**Supplementary Figure 2A:**
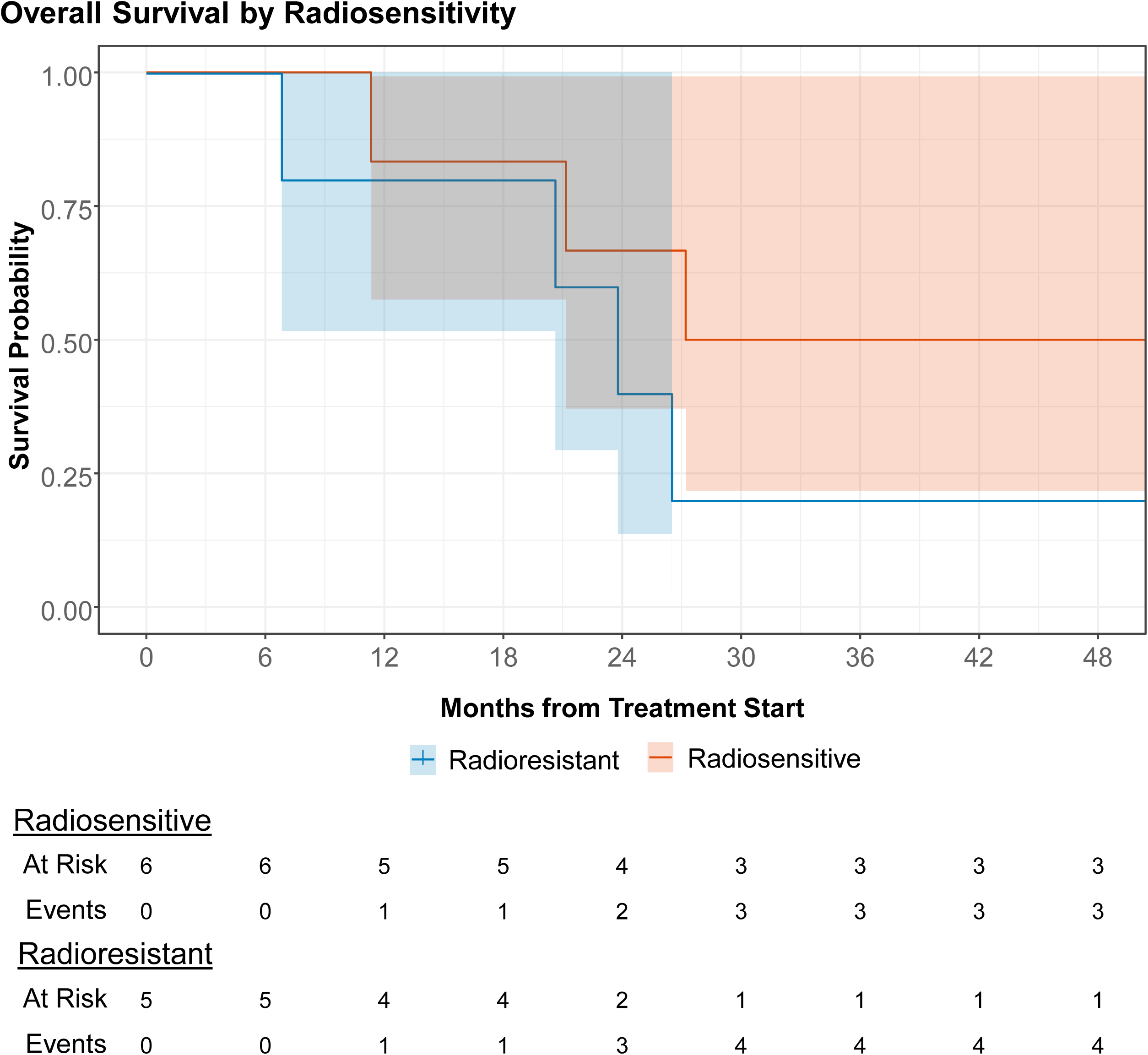
Overall Survival by Radiosensitivity. Radioresistant tumoroids (n=5) had a lower overall survival in the studied time period than the radiosensitive tumoroids (n=6), though sample size precludes statistically significant comparisons of these groups.

**Supplementary Figure 2B:**
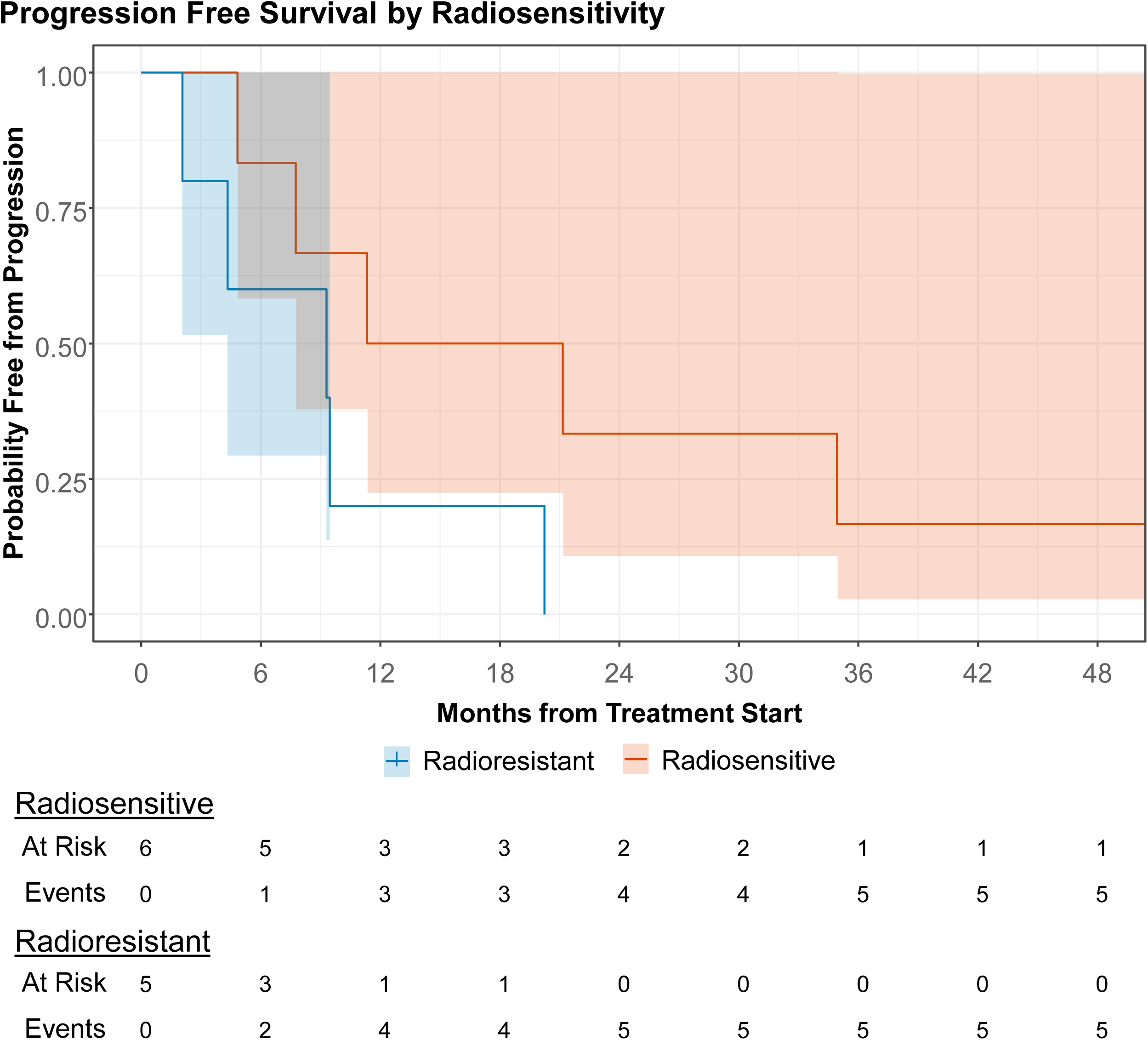
Progression-Free Survival by Radiosensitivity. Radioresistant tumoroids (n=5) had a lower progression free survival in the studied time period than the radiosensitive tumoroids (n=6), though sample size precludes statistically significant comparisons of these groups.

**Supplementary Figure 3:**
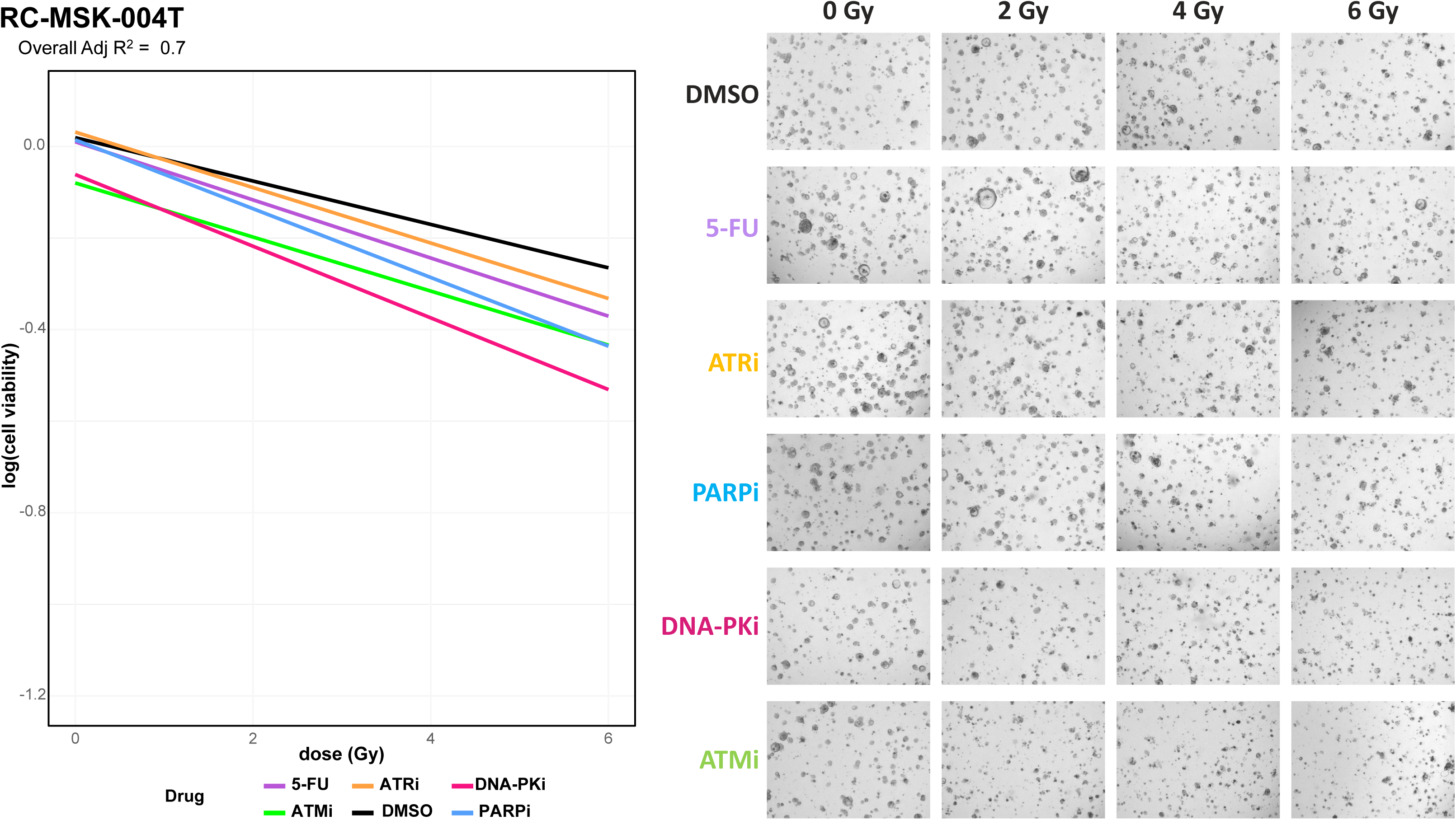
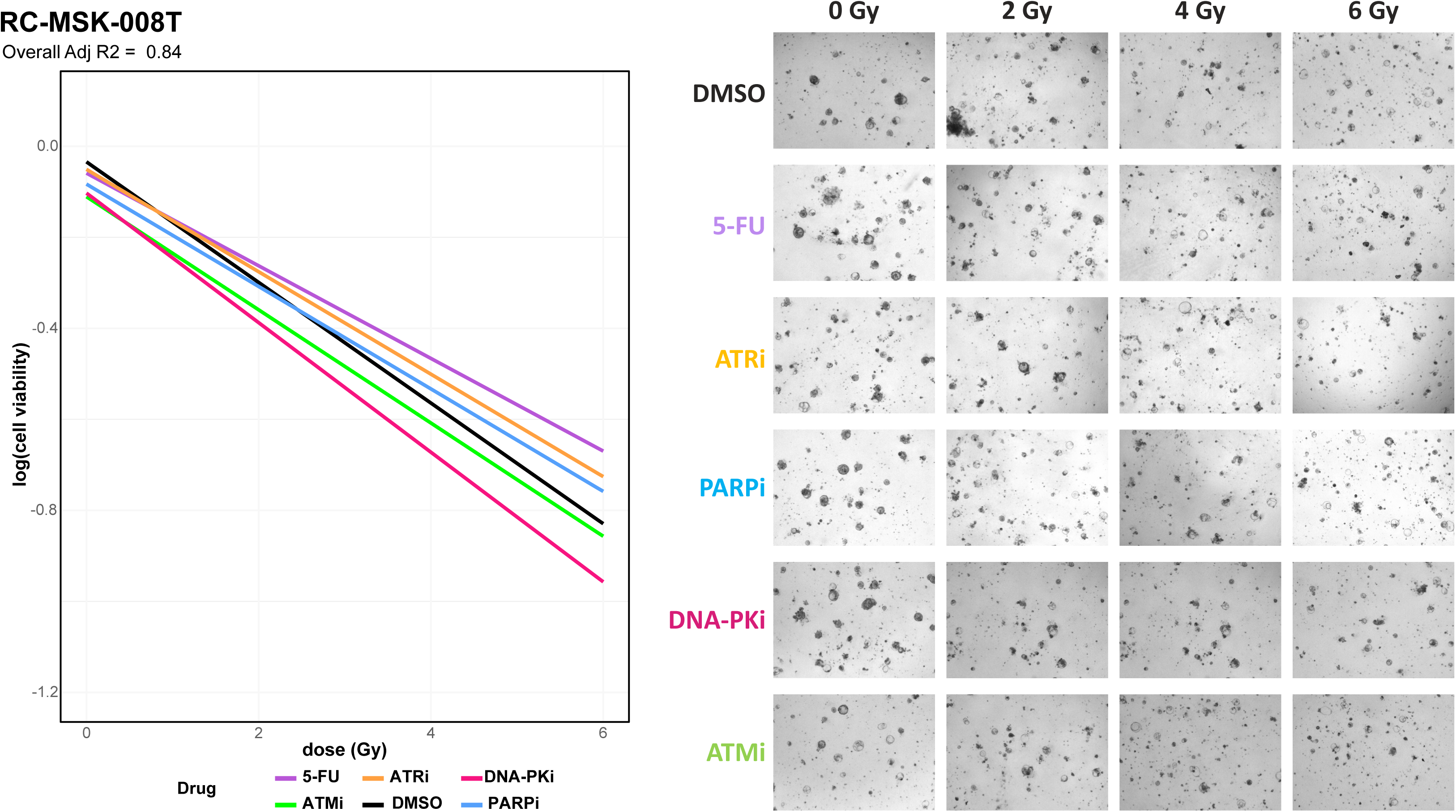
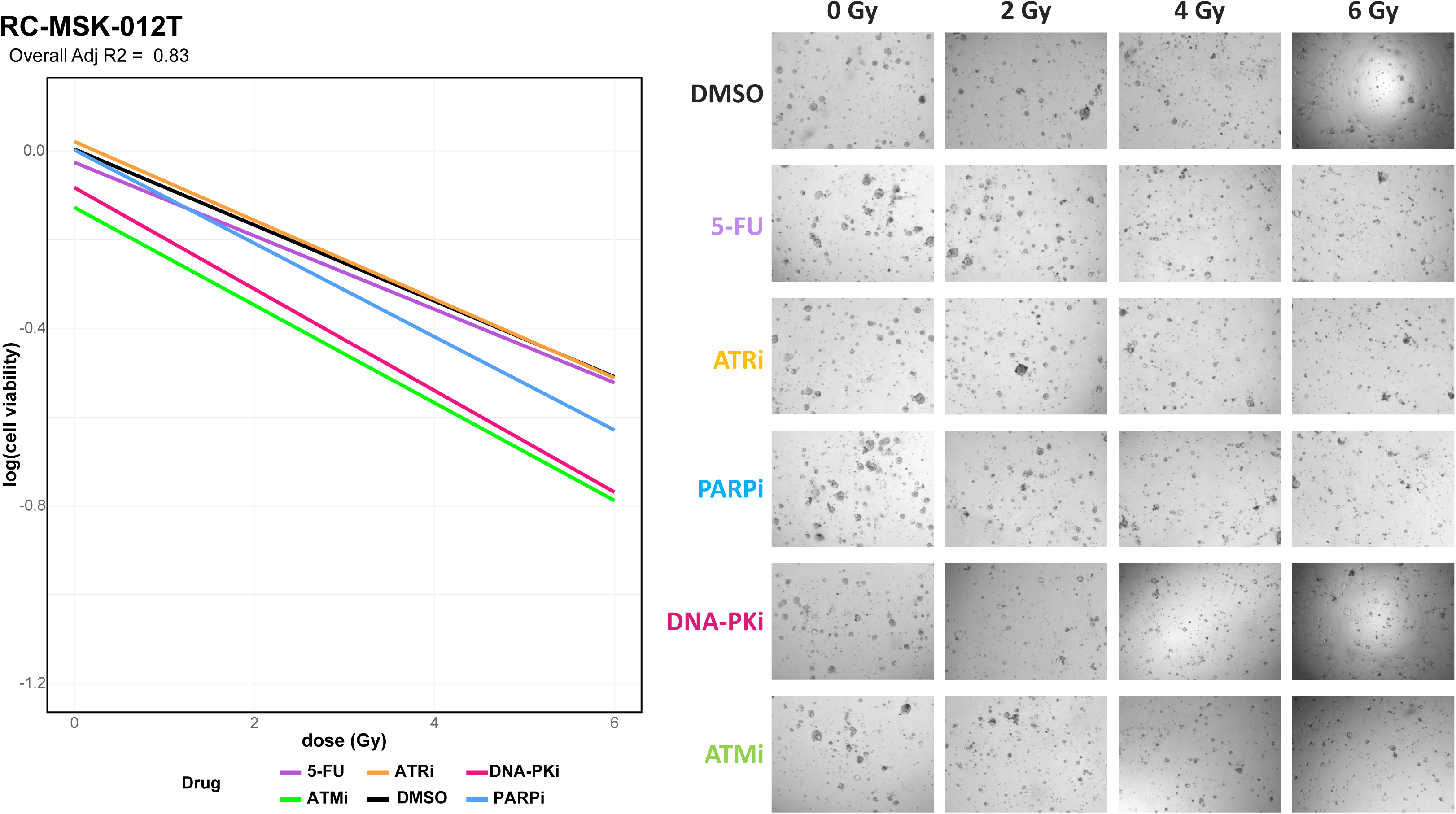
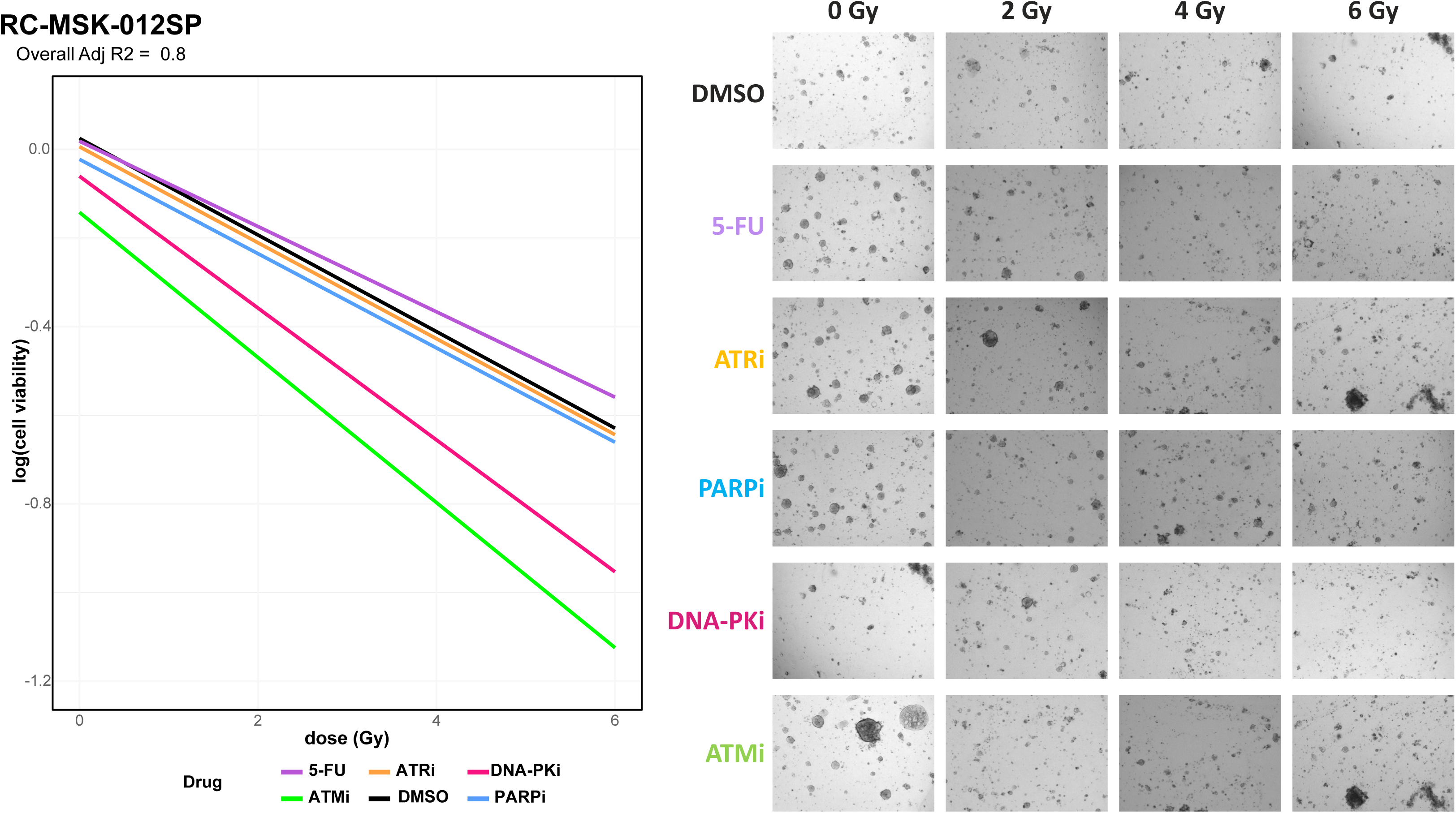
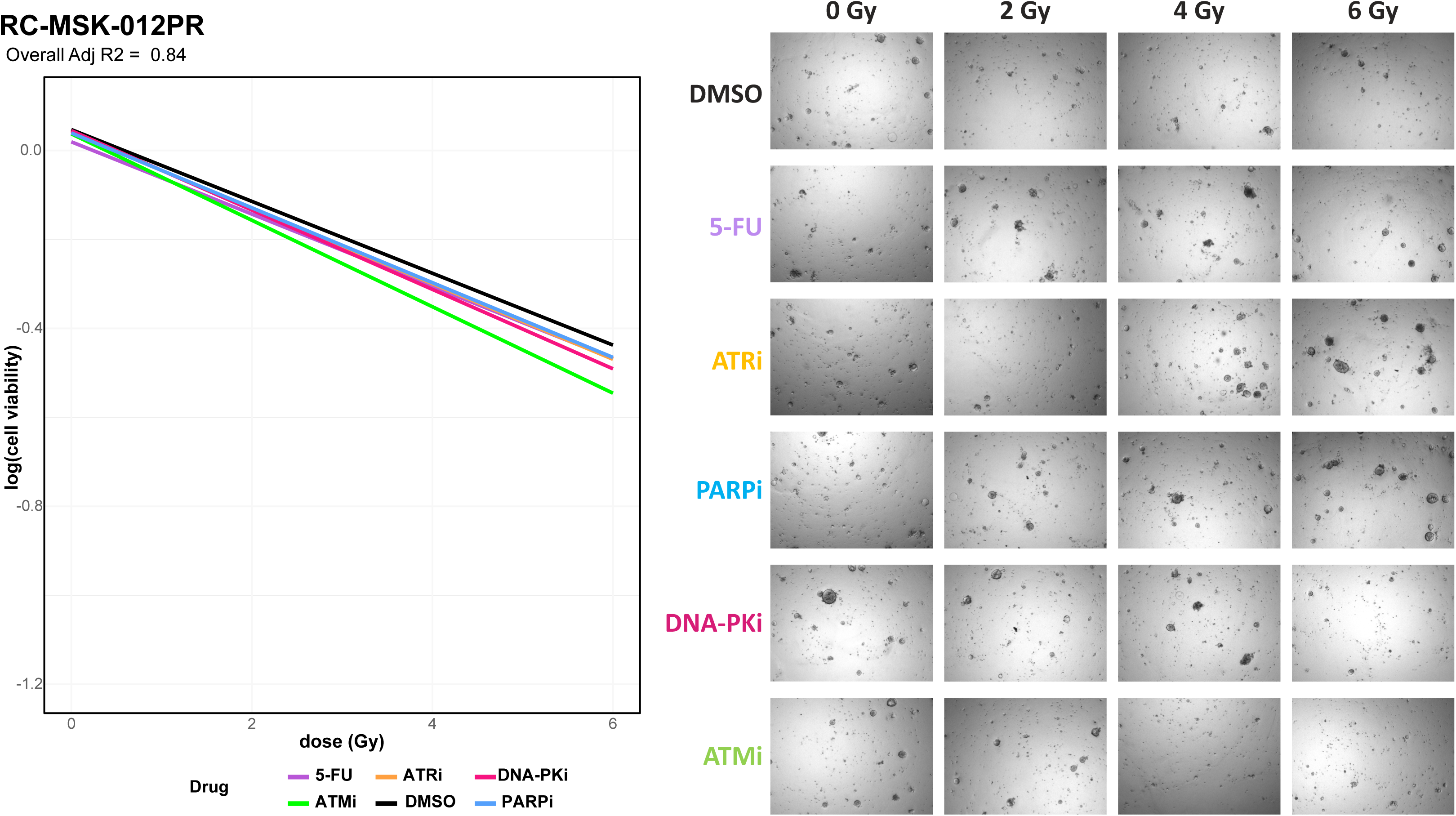
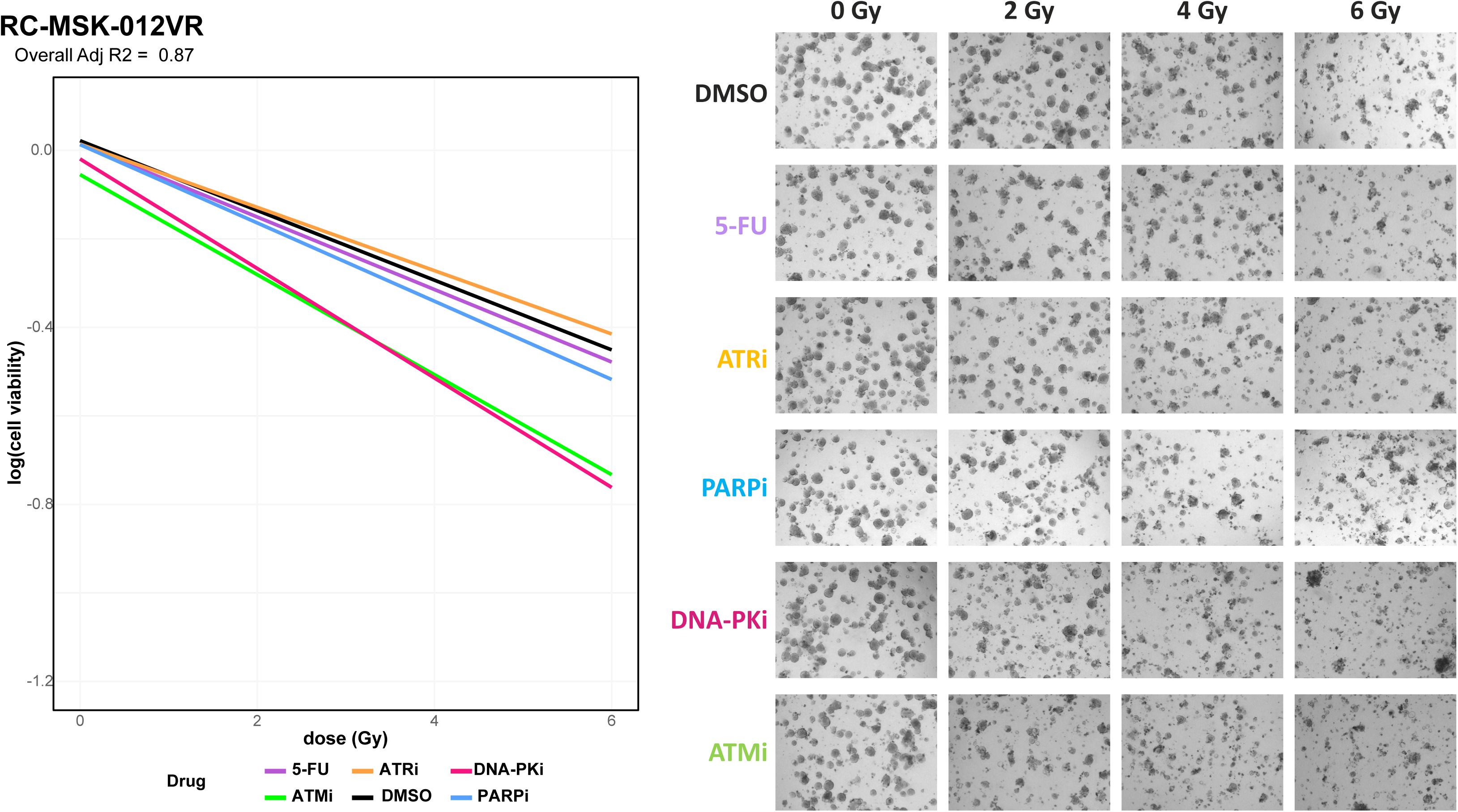
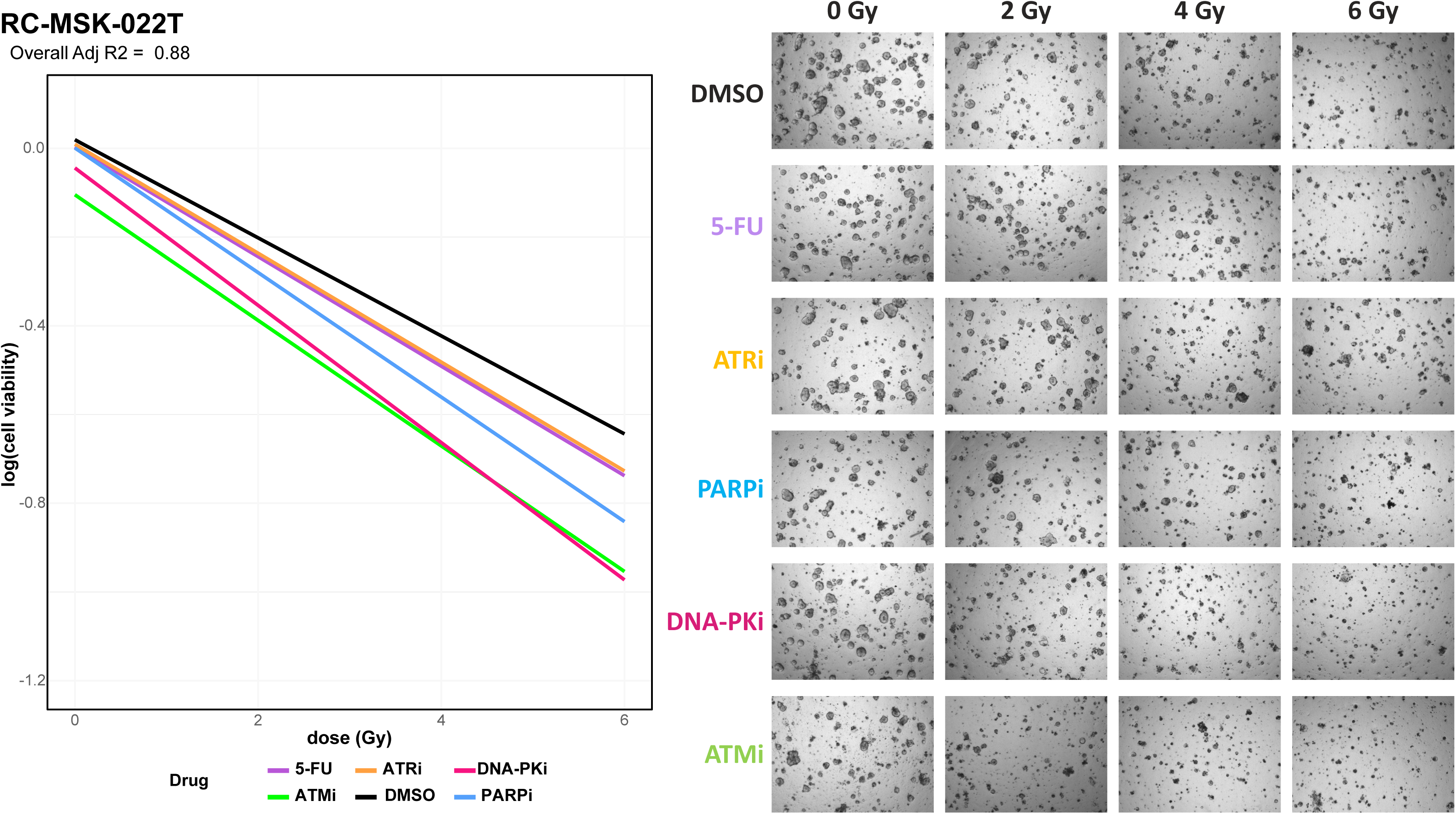
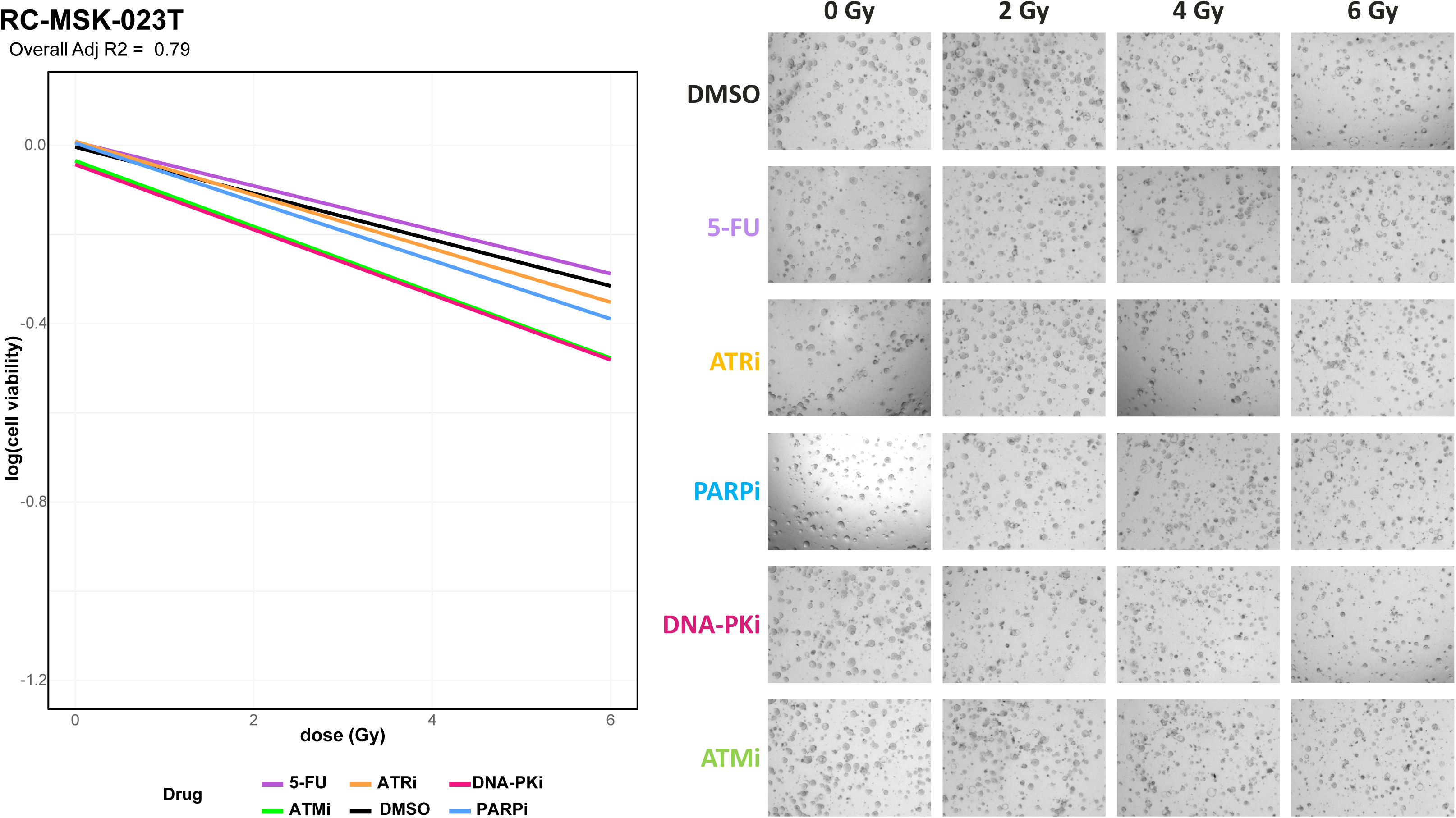
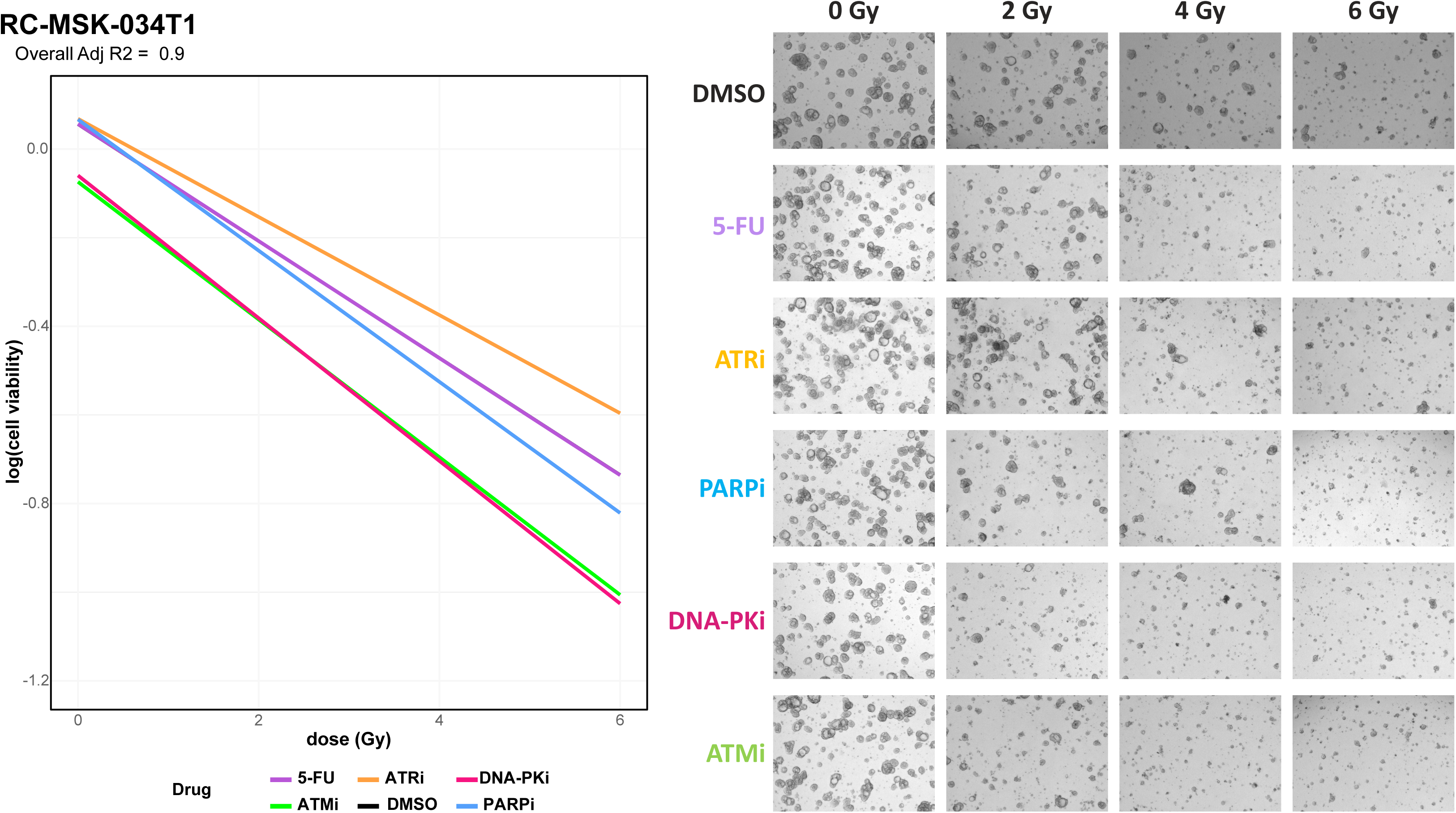
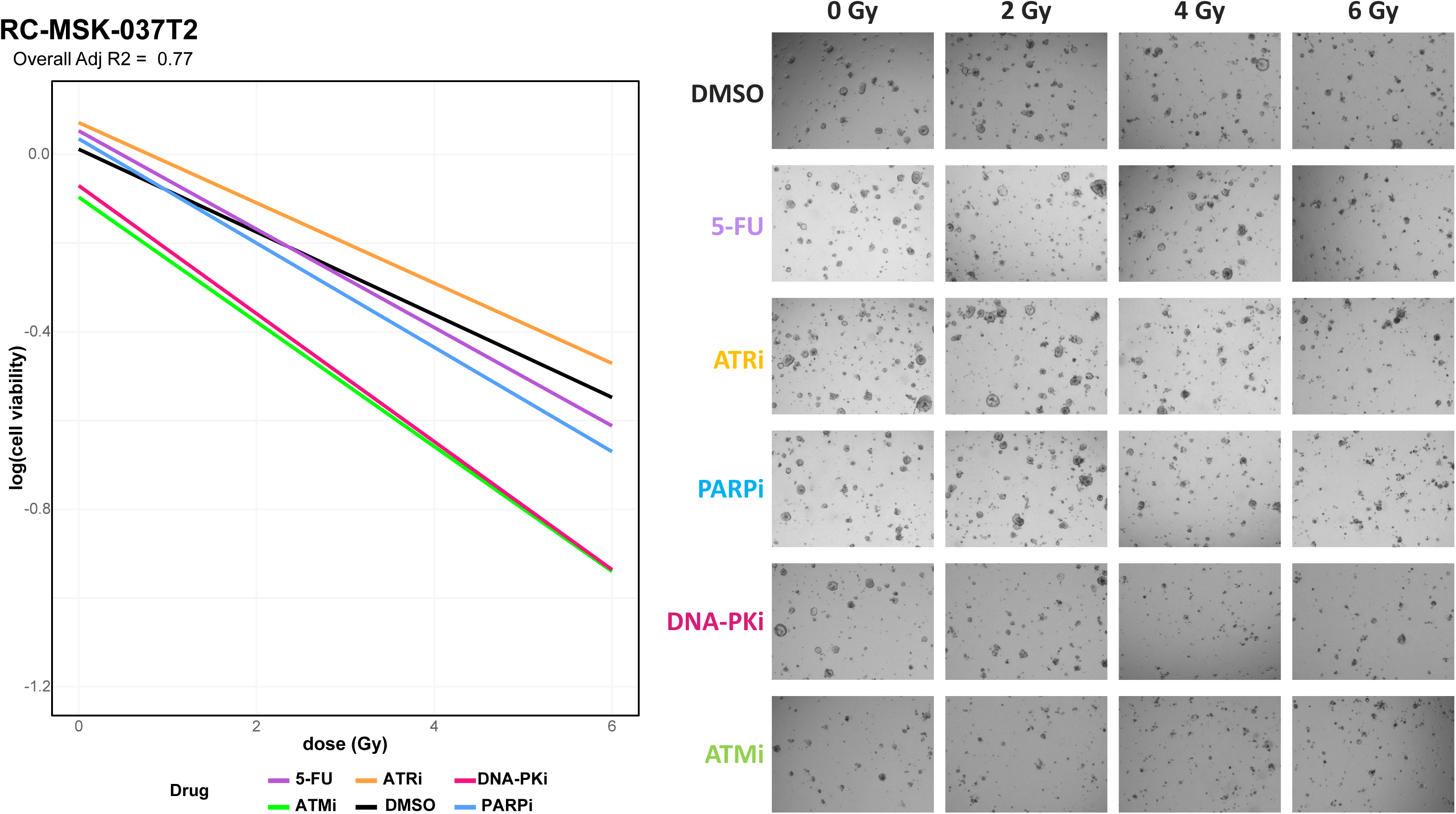
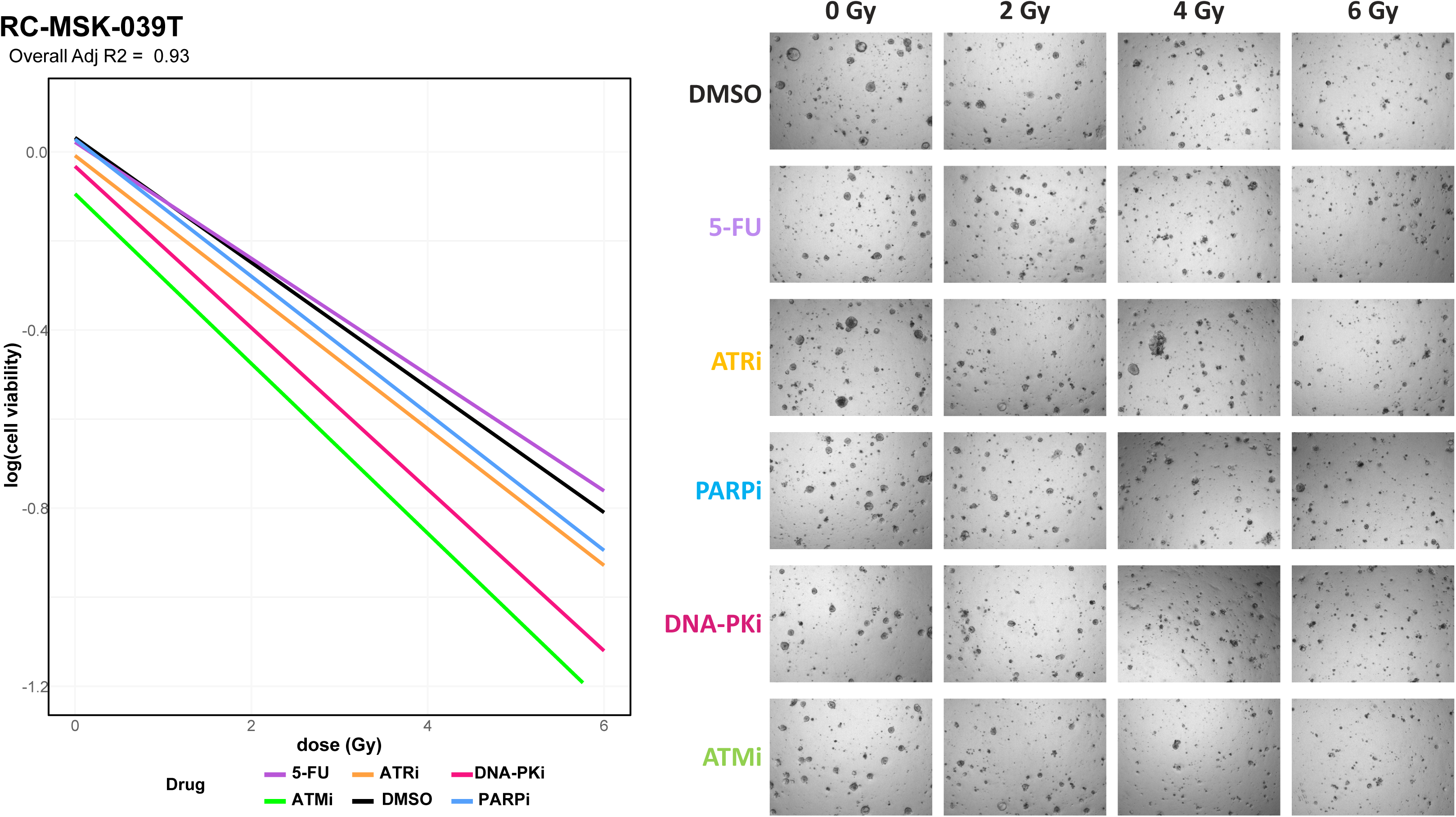
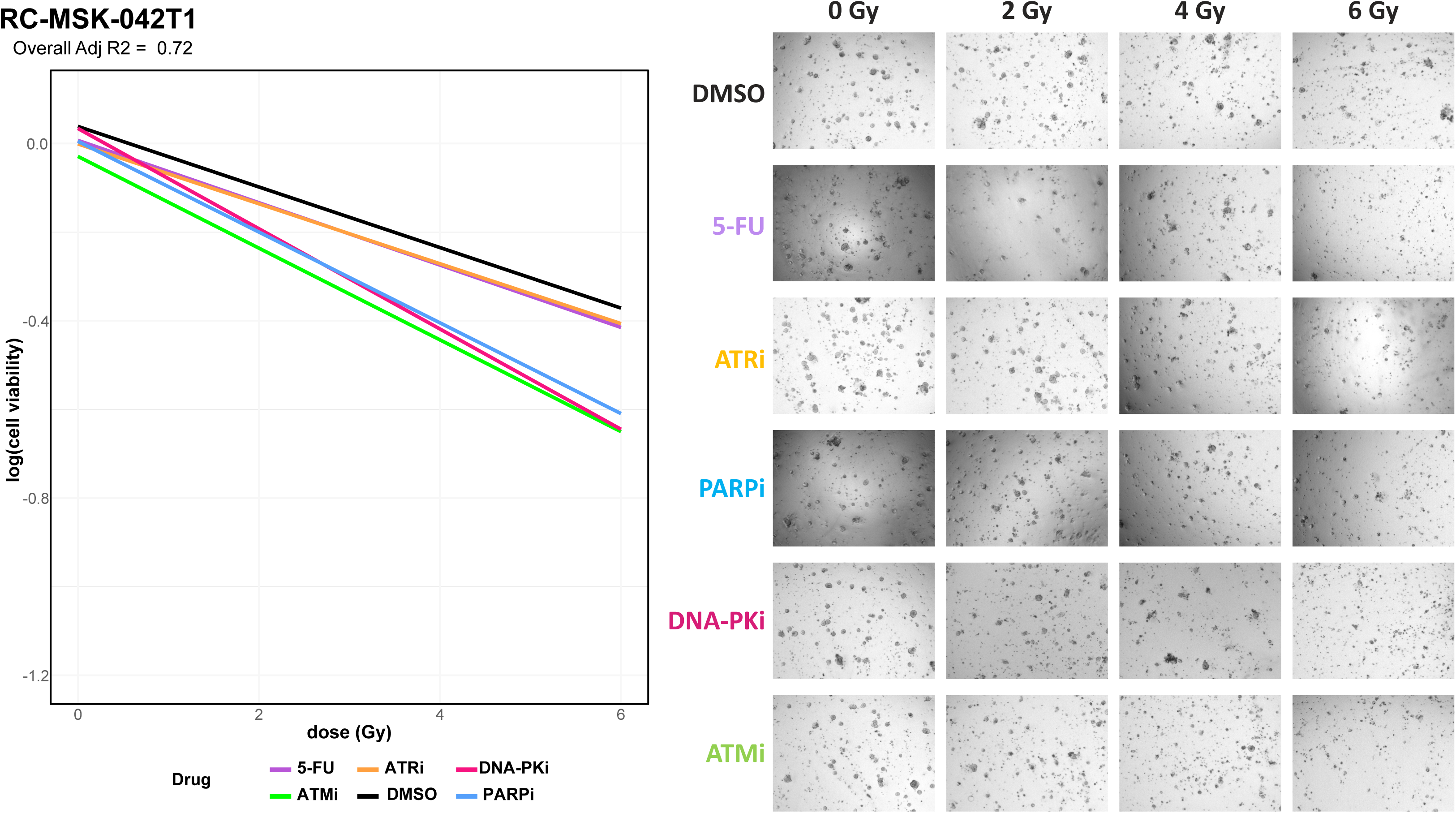
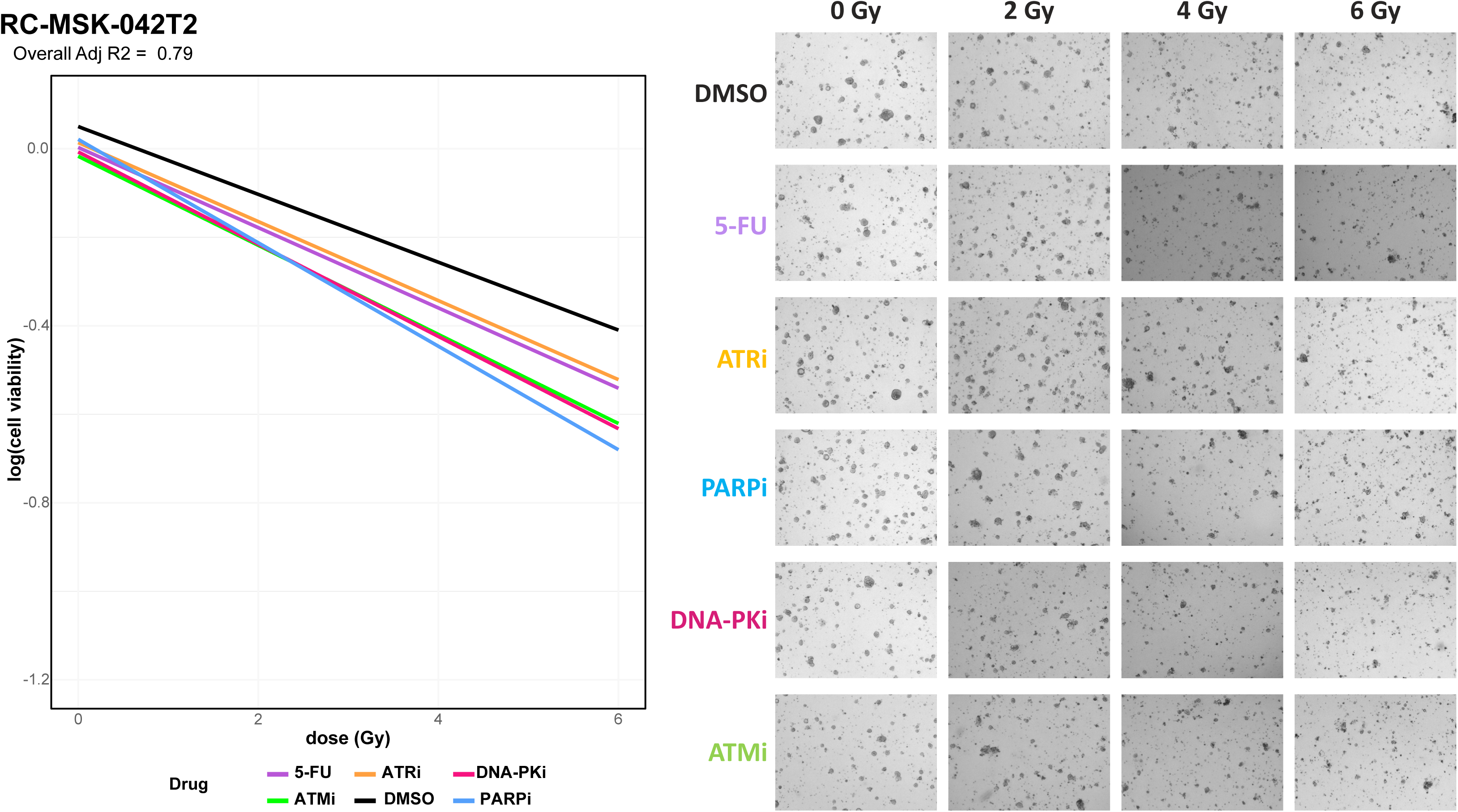
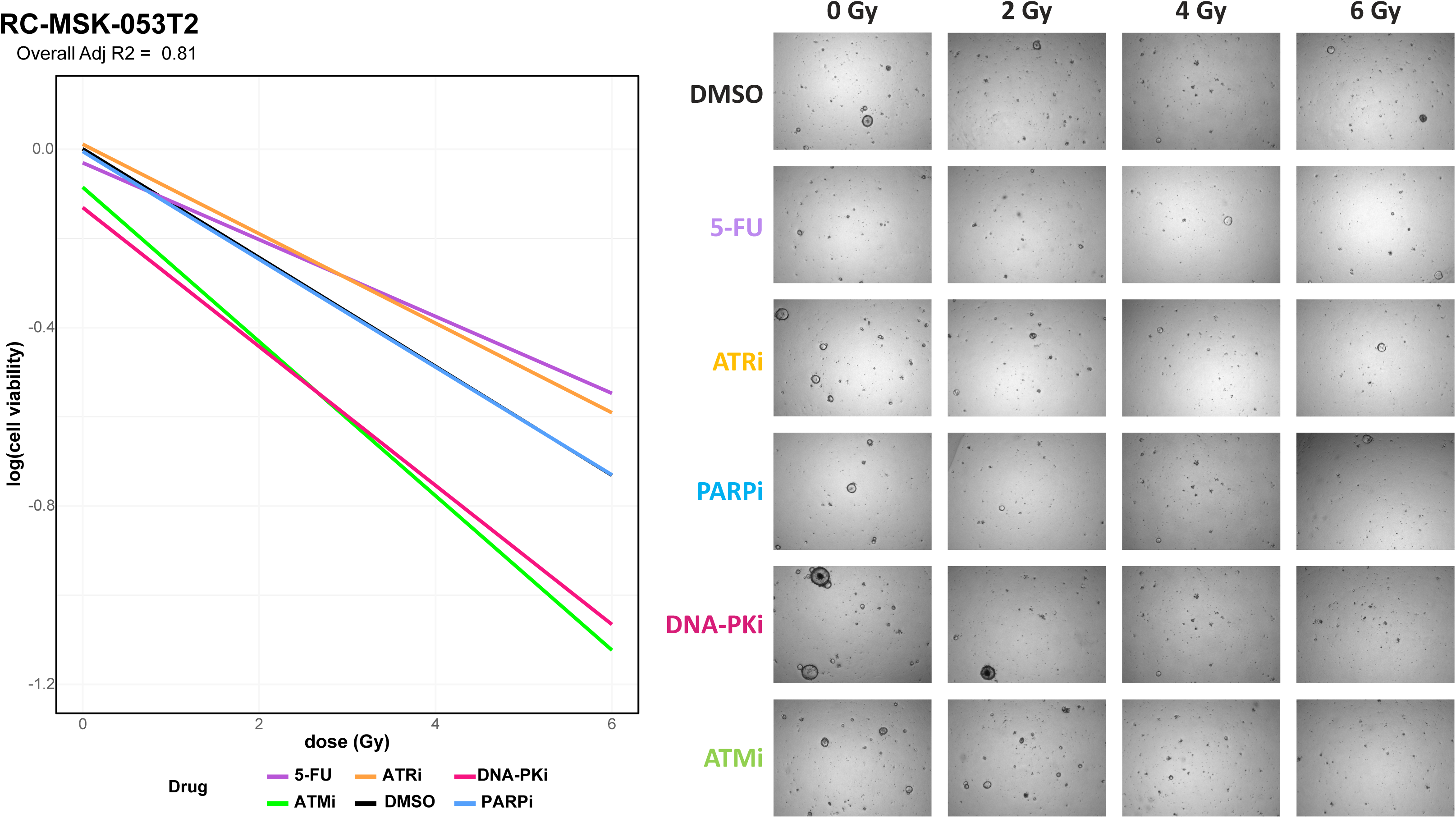
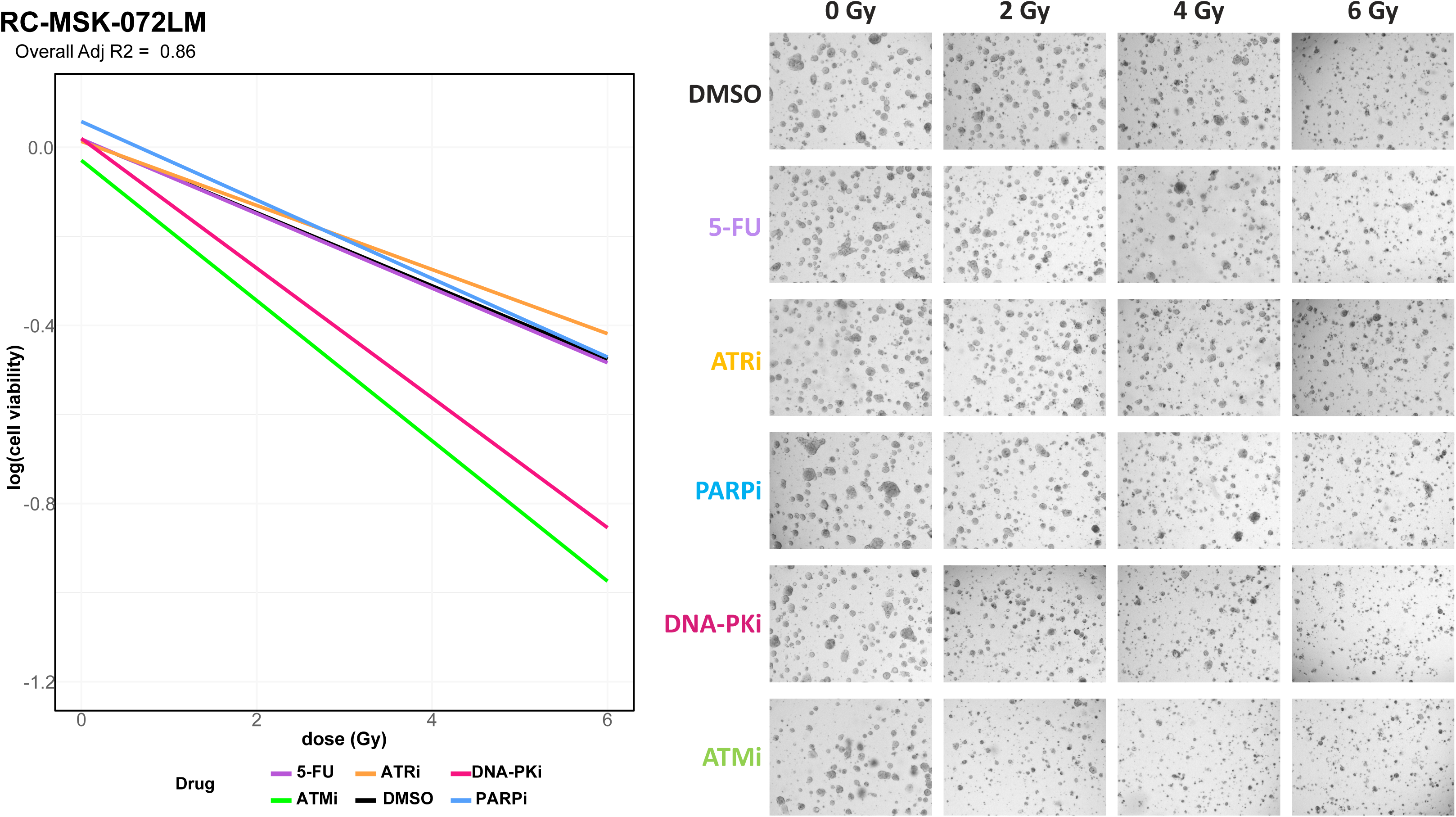
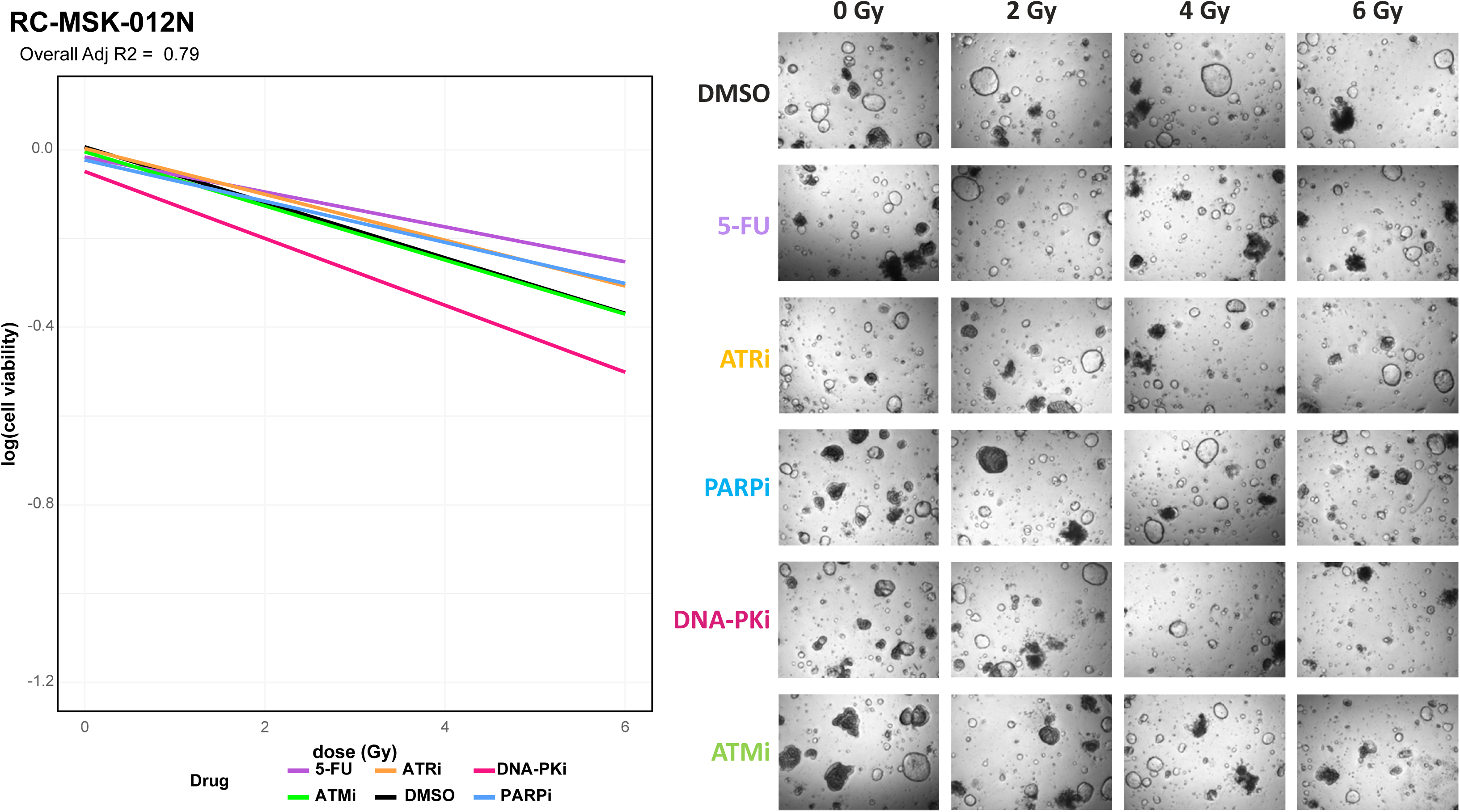
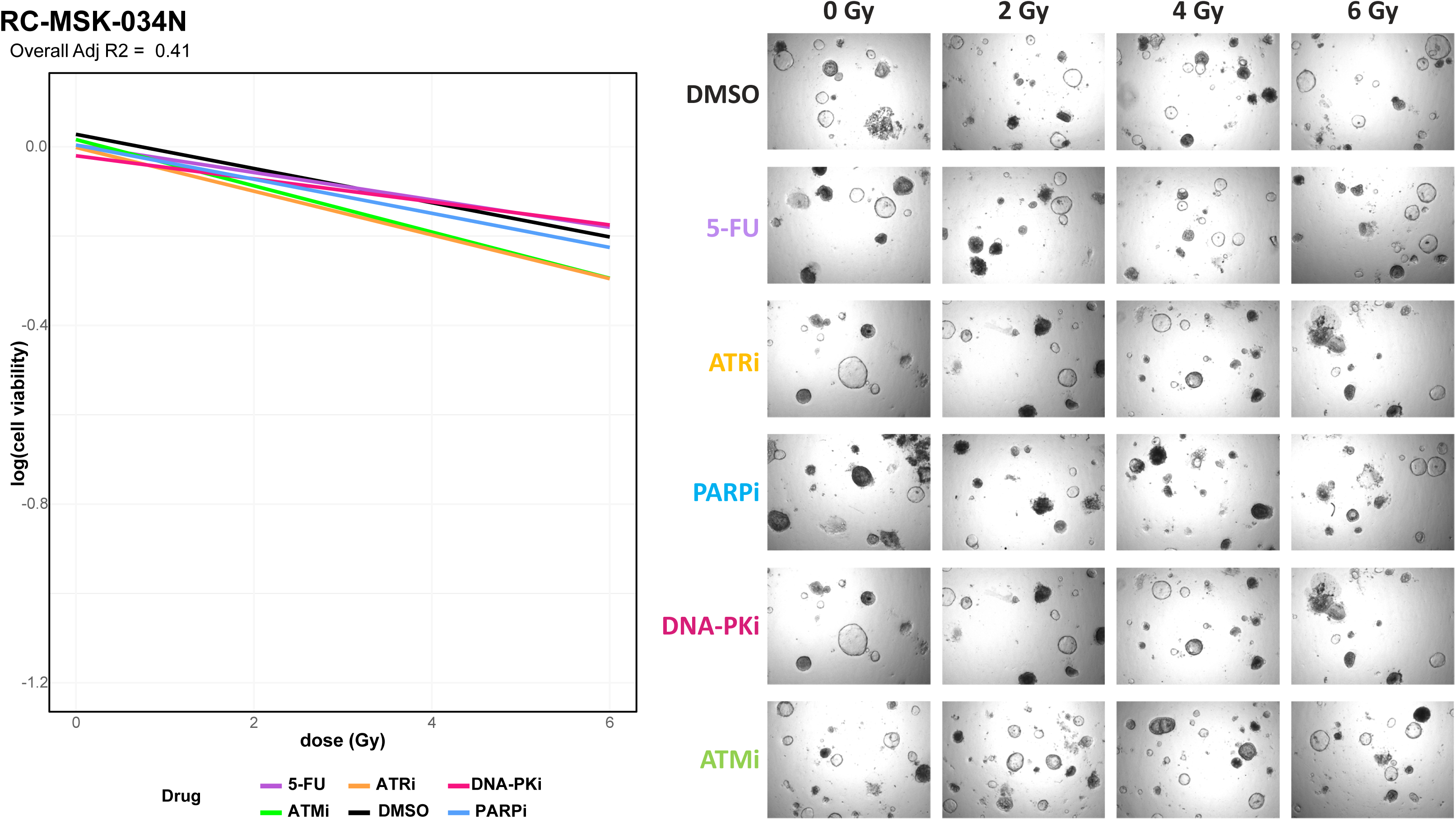
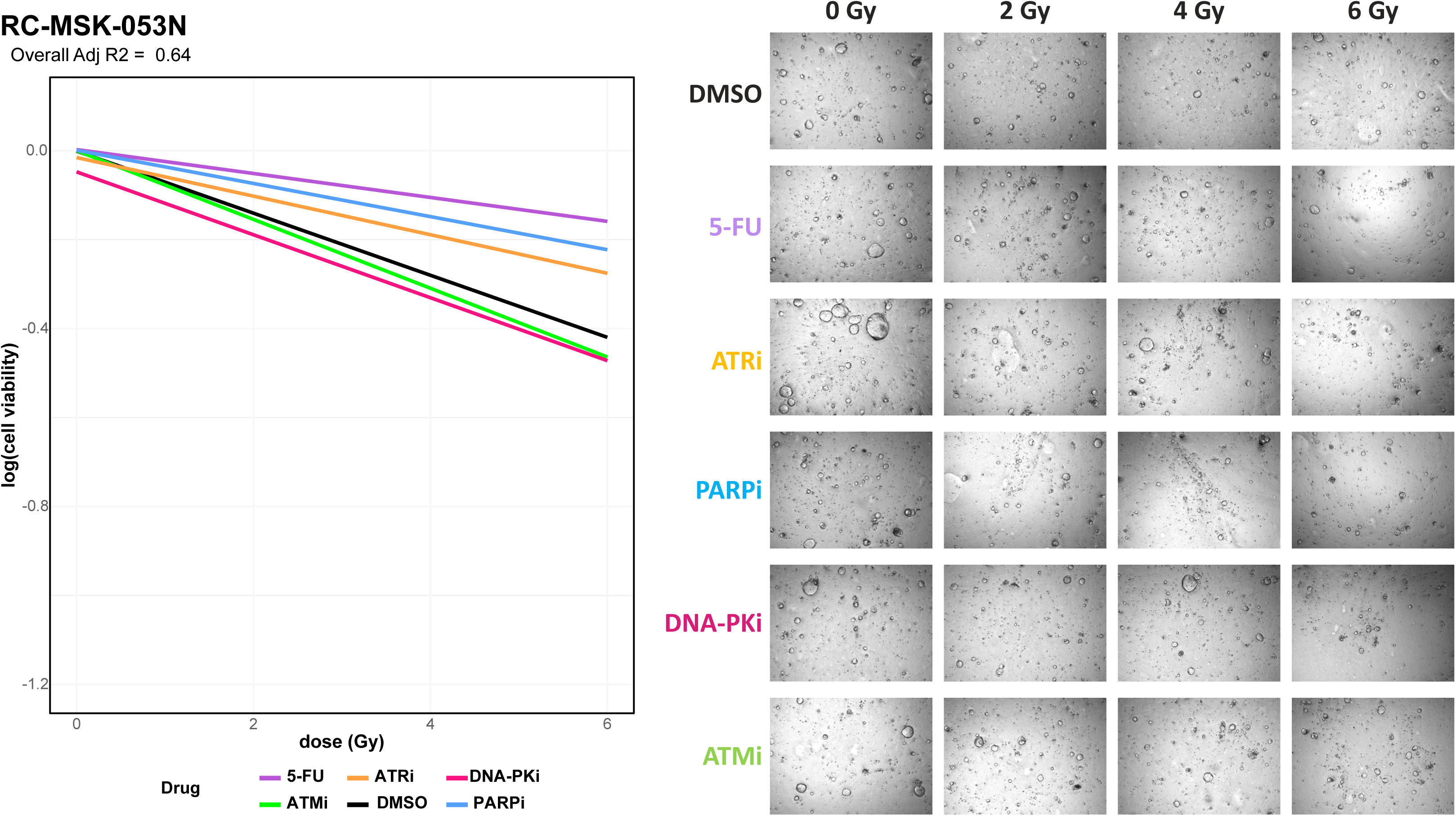
Linear Cell Viability Plots for Tumoroids and Organoids. Cell viability plots demonstrate change in log-transformed linear cell viability as radiation dose increases. Drug treatment conditions can be seen in corresponding colors. Corresponding brightfield microscopy images are shown with each plot.

**Supplementary Figure 4A-4D:**
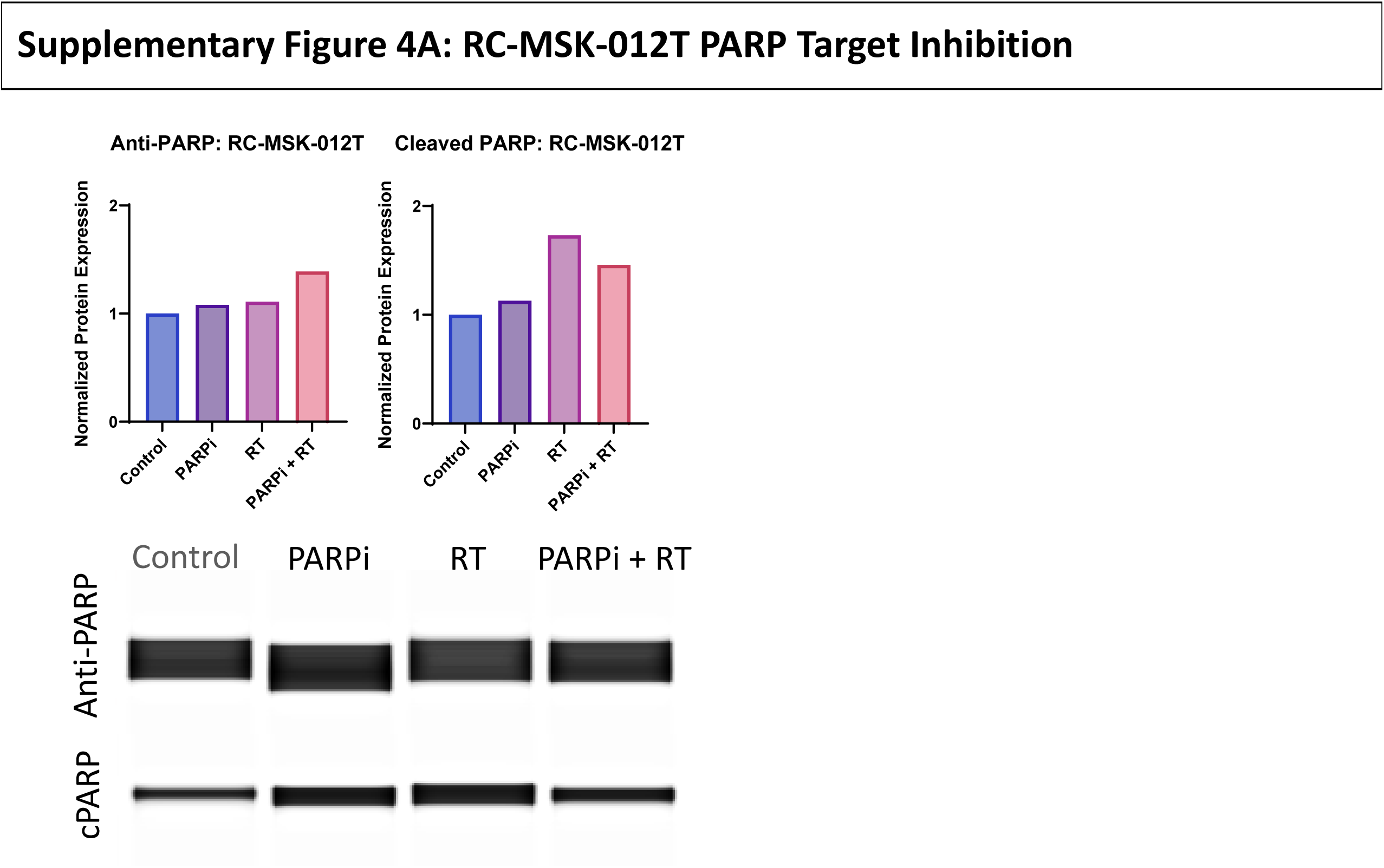
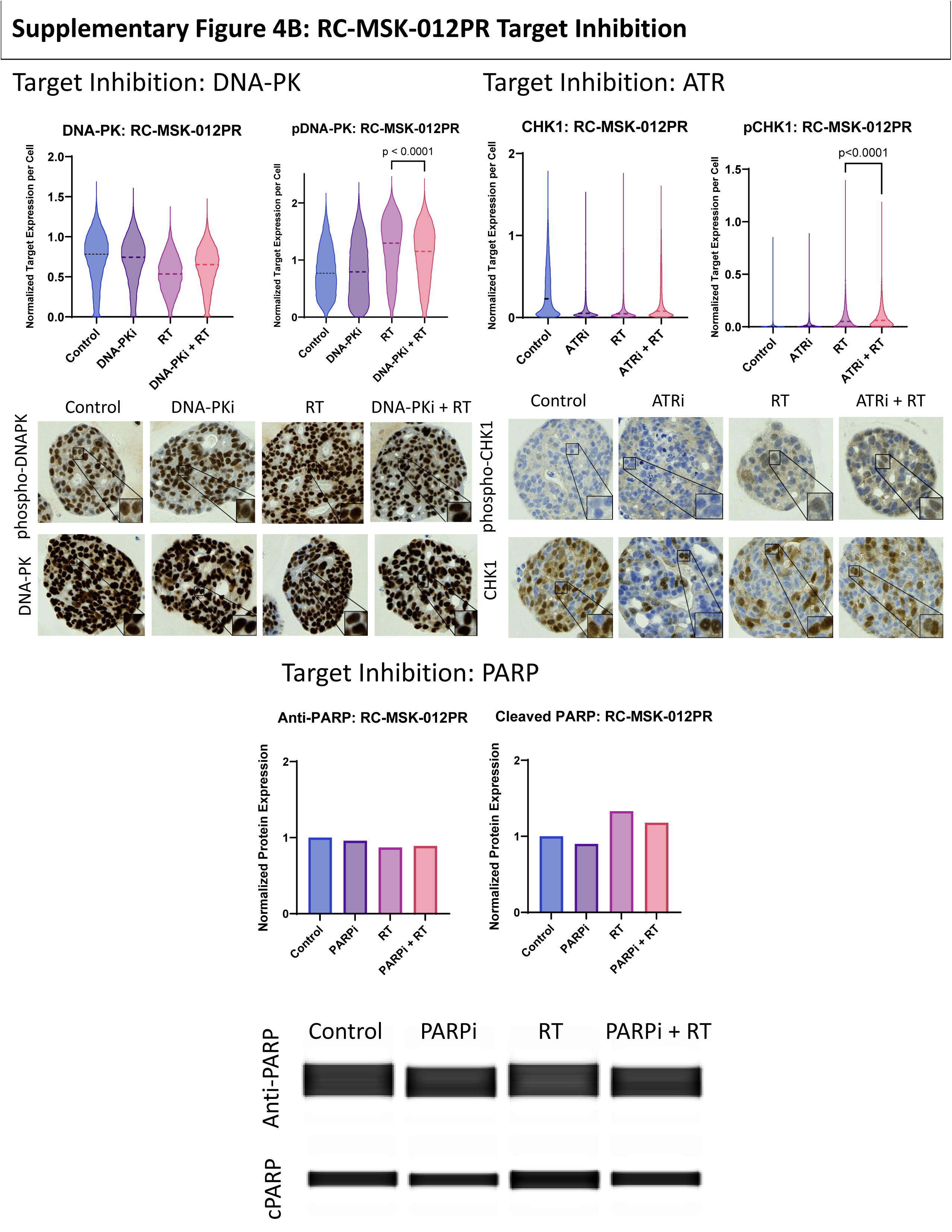
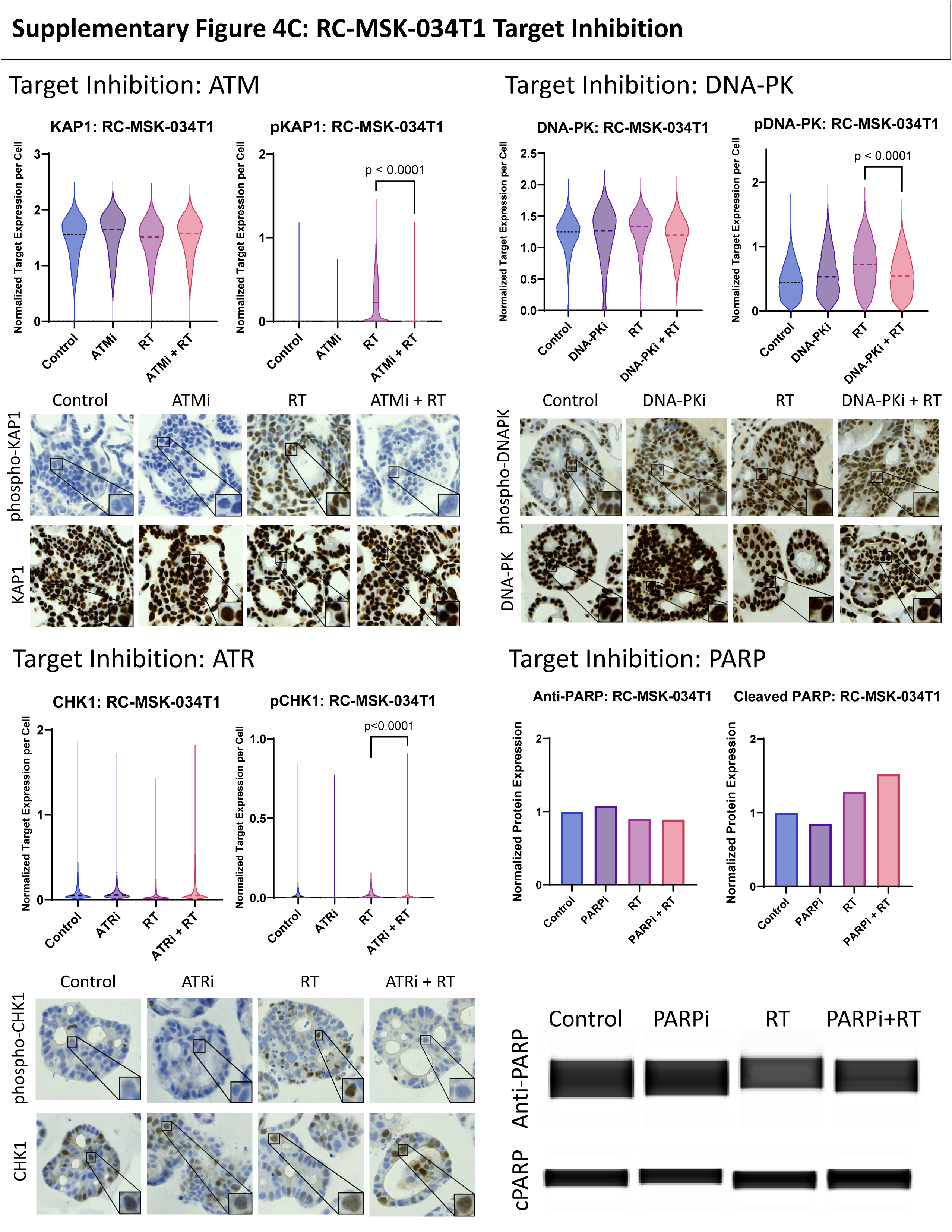
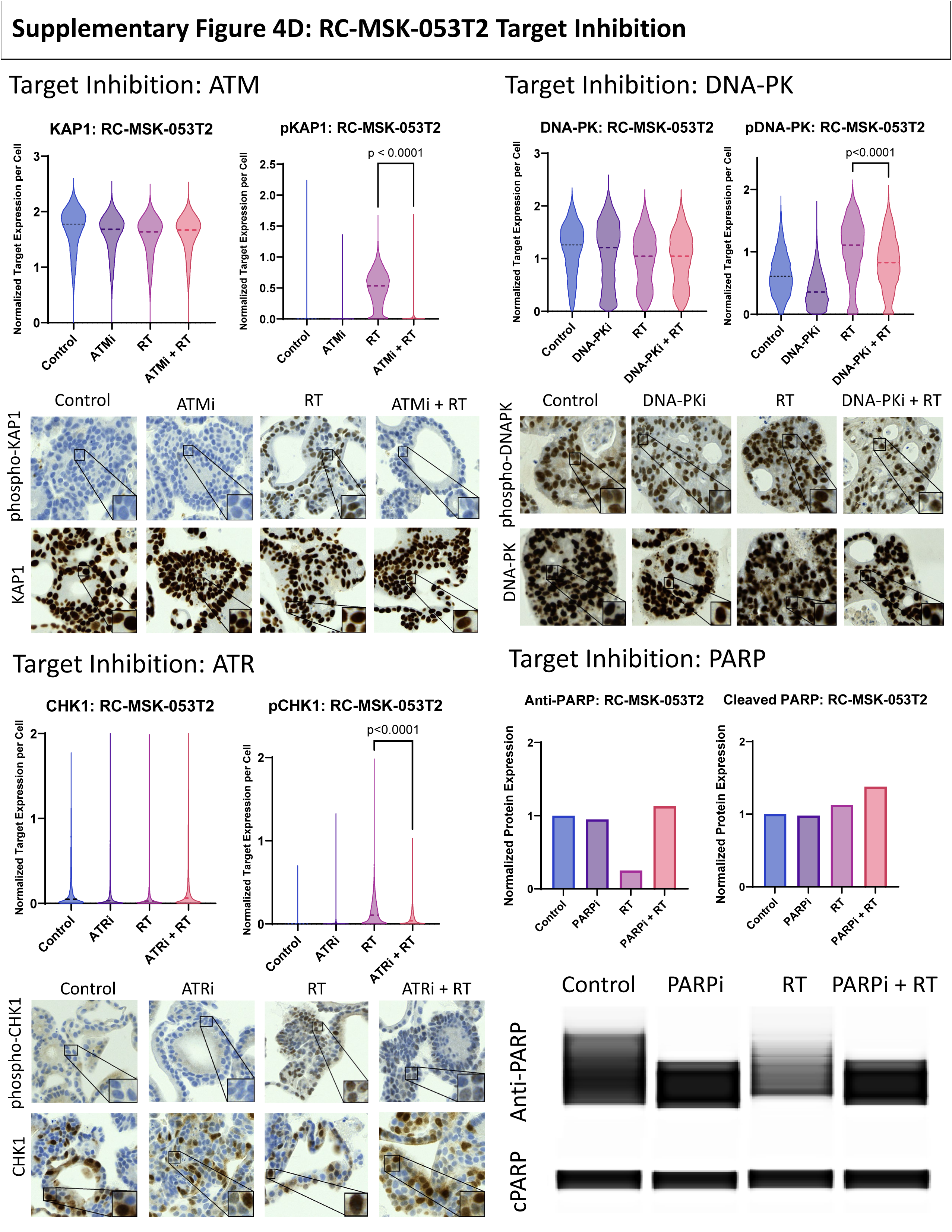
Target Inhibition for RC-MSK-012PT, RC-MSK-012PR, RC-MSK-034T1, and RC-MSK-053T2. Inhibition of ATM was demonstrated by staining of downstream target KAP1 and phospho-KAP1. Inhibition of DNA-PK was demonstrated by staining of DNA-PK and phospho-DNAPK. Inhibition of ATR was demonstrated by staining of downstream target CHK1 and phospho-CHK1. When treated with radiation (RT), staining demonstrates phosphorylation of target proteins. However, when treated with inhibitor and RT, downstream phosphorylation is inhibited. Inhibition of PARPi was demonstrated using the JESS Automated Western Blot system per kit protocol. RT demonstrates an increase in cleaved PARP (cPARP), whereas treatment with RT and inhibitor demonstrates reduced levels of cleaved PARP.

**Supplementary Figure 5:**
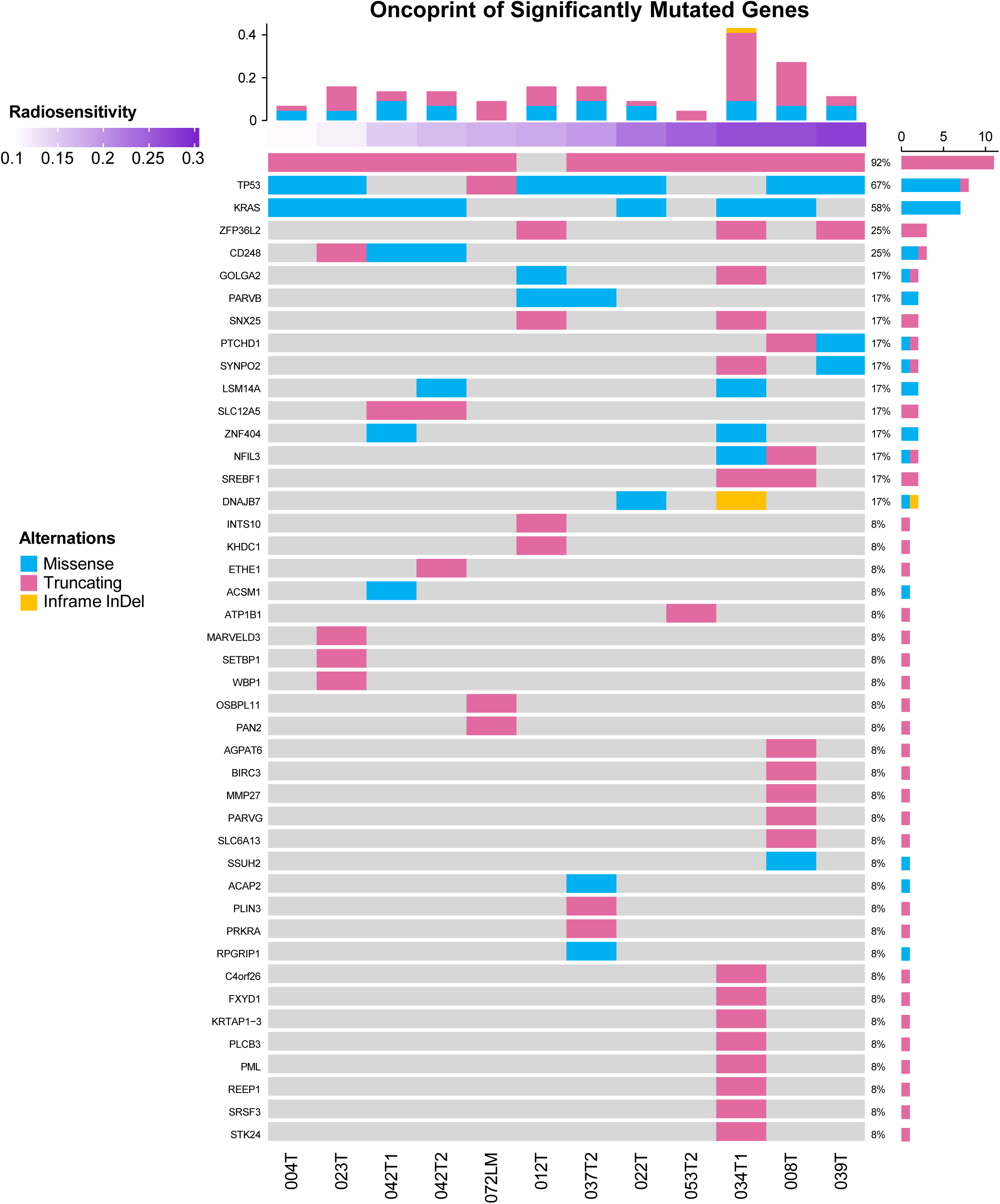
Among the 44 significantly mutated genes (MutSig2CV p-value < 0.01), the most frequent mutations included APC present in 92% of patients, TP53 (67% of patients), and KRAS (58% of patients). Despite the focus on radiation sensitivity, no somatic mutations were detected in DNA damage repair (DDR) pathway genes.

## Notes

### Author Declarations

The Memorial Sloan Kettering Cancer Center gave ethical approval for this work under the existing protocol 16-1071.

