## Supplemental Methods for "Leveraging a Patient-Derived Tumoroid Platform for Precision Radiotherapy: Uncovering DNA Damage Repair Inhibitor-Mediated Radiosensitization and Therapeutic Resistance in Rectal Cancer"

**Supplementary Methods**

*Normal Organoid Derivation*

Normal organoids were derived from adjacent healthy rectal tissue in a process similar to tumoroid derivation but with some key modifications.[7] A biopsy of normal rectal tissue was taken at least 4-10 cm away from/proximal to the tumor and immediately transferred to ice-cold PBS. Several steps were subsequently required to isolate crypts from the normal tissue. First, the specimen was gently rinsed with 1 mL of ice-cold PBS-Abs buffer, transferred to a 1.5 mL sterilized Eppendorf tube, and further rinsed with 0.5 mL of ice-cold EDTA-PBS (0.5 M EDTA solution was diluted in PBS to obtain an 8 mM final concentration of EDTA). Next, the EDTA-PBS was removed manually, and another 1 mL of ice-cold EDTA-PBS was added to the Eppendorf tube. The biopsy sample was confirmed to flow freely within the tube, and then the tube was rolled gently on a roller for 60-70 minutes at 4^o^C. We then proceeded to shake the Eppendorf tube vigorously by hand until small particles were shaken loose from the specimen and appeared floating in the buffer. Under a microscope, the tiny particles were confirmed to be fragments of crypts. We then moved to transfer the buffer with particles (crypts) to a new 15 ml conical tube, taking care to not disturb the specimen. Another 1 mL of ice-cold PBS was added to this Eppendorf tube with the specimen, and the manual crypt isolation by vigorous shaking was repeated 7-10 times to ensure high yield of crypts from the normal specimen. All buffer with particles (crypts) was transferred to the same 15 ml conical tube and subsequently centrifuged at 500×g for 5 minutes. The supernatant was carefully removed, and the remaining pellet was resuspended in 0.5 mL of TrypLE Express. The tube was incubated for 15 minutes at 37^o^C. Then, the mixture was diluted with 10 mL of ice-cold PBS and again centrifuged at 500×g for 5 minutes. The supernatant was carefully removed, and the resulting pellet was resuspended in 300 μL of Matrigel. The Matrigel mixture was seeded as 40 μl domes in a 24-well cell suspension plate and solidified in the tissue culture incubator. Approximately 800 μL of complete medium was added to the wells with the Matrigel domes, which were then cultured in regular tissue culture conditions to form organoids. The organoids were maintained and expanded in the full medium, and the culture medium was refreshed every 2-3 days. Full medium was prepared as follows: advanced DMEM/F12 was supplemented with 1×antibiotic-antimycotic (Gibco), 1×B27, 1×N2 (Gibco), 1×GlutaMAX, 10 nM gastrin I, 10 mM HEPES, 1 mM N-acetylcysteine, 10 mM nicotinamide, 100 ng/mL human recombinant EGF, 500 nM A83–01, 10 μM SB 202190, 10 μMY-27632, and 50% L-WRN conditioned medium, which was derived from L-WRN cell (ATCC CRL3276) producing L-Wnt3A, R-Spondin-3, and Noggin. In contrast, tumor medium is prepared without the 50% L-WRN conditioned medium.

*Radiosensitizer Experiment*

Tumoroids were seeded in a clear-bottomed 96-well plate. To prepare the plate, each well in the plate was coated with 200 μl of an ice-cold 12% Matrigel in sterilized water, centrifuged at 500xG at 4°C for 5 minutes, and incubated overnight to solidify Matrigel layer.

The next day (Day 1), the sterilized water was removed from each well, leaving behind a Matrigel layer. Then, the tumoroid line of interest was trypsinized and cells dissociated. The tumoroid cells were resuspended in a solution of tumor media and 5% Matrigel, with enough cells to achieve 3000 cells per well. Exactly 100 μl of the tumoroid-media suspension was added to each well, followed by centrifugation and incubation to solidfy the Matrigel. Two hours later, 150 μl of media spiked with drug solution was added to the tumoroid-seeded wells. See Supplementary Table 1 for a list of DDRi drugs and concentrations used. After two hours of drug treatment, the tumoroids were irradiated at doses of 0 Gy, 2 Gy, 4 Gy, or 6 Gy using an XRad 320 Irradiator. The tumoroids were incubated for four more days with replenishing of the drugged medium on Day 5.

On Day 9 of the experiment, the drugged medium was carefully aspirated without disturbing the Matrigel layer. The CellTiter-Glo 2.0 reagent (Promega, Catalogue #G9241) was mixed with PBS in a 1:1 ratio and 200 μl was added to each well, pipetting gently to disrupt the Matrigel layer. Using a luminometer (Promega Glomax), the number of viable cells was measured for each drug and radiation condition. Experiments were conducted in technical quadruplicate and biologic triplicate or quadruplicate.

The procedure for normal organoids was identical but used complete medium instead of tumor medium, with pilot studies demonstrating that medium choice did not affect drug efficacy.

*Pilot Radiosensitizer Experiments:*

Prior to conducting the above radiosensitizer experiment, pilot studies were required to determine appropriate dosages for DDRis for optimal effect without toxicity. Additionally, the effects of different mediums on drug efficacy were tested as well. Two tumoroids (RC-MSK-023T and RC-MSK-039T) were used for the pilot studies. Media conditions included tumor medium or complete medium. Drug concentrations included the following: for ATMi and DNA-PKi, dosages tested included 0 μM, 0.05 μM, 0.1 μM, 0.5 μM, 1 μM, and 5 μM. For ATRi, dosages tested included 0 μM, 0.01 μM, 0.02 μM, 0.1 μM, 0.2 μM, and 1 μM. PARPi was previously found to have minimal toxicity at 5 μM.

Tumoroids were plated and incubated as described above, but with differing media conditions and varying drug concentrations. Drugged media was replenished on Day 5, and cell viability was measured using the CellTiter-Glo assay on Day 9, as above.

Cell viability in each condition was plotted over the increasing dose of agent. It was found that dose response curves did not differ greatly between different medium conditions. Drug concentrations were selected for the experiment such that decreases in cell viability were approximately 10% at the specified dose, minimizing toxicity from the DDRi and ensuring that the effects of radiation and sensitization were minimally impacted.

*Target Inhibition by Immunohistochemistry*

Inhibition of drug targets or downstream targets for ATM, DNA-PK, and ATR was observed by immunohistochemistry. Phospho-KAP1 was used to characterize ATM activity, phospho-DNA-PK was used to characterize DNA-PK activity, and phospho-CHK1 was used to characterize ATR activity. The basal expression of ATM, DNA-PK, and ATR in each condition was also measured as a control. Target inhibition was initially validated in three tumoroids (RC-MSK-012T, RC-MSK-034T1, and RC-MSK-053T2), then expanded to include a post-progression tumoroid (RC-MSK-012PR).

Tumoroids in all treatment conditions (no treatment, drug only, radiation only, combination drug and radiation) were fixed in 10% buffered formalin phosphate (Fisher SF100-4). For each inhibitor, fixation was done at several time points after radiation to capture greatest DDR protein and inhibitor activity. Fixed tumoroids were incubated in 70% ethanol until paraffin embedding by standard technique. Slides were cut from FFPE blocks for subsequent immunohistochemistry. Slides were baked at 64°C for 1 hour, then transferred to Leica Bond Rx for staining. Antigen retrieval was performed with Leica Epitope Retrieval Solution 2 (AR9640) for 30 minutes at 100°C. Antibodies were then incubated for 30 minutes. Supplementary Table 2 displays antibodies and dilutions used for each DDRi. Antibodies were then stained using the Leica Bond Polymer Refine Detection Kit (DS9800). Slides were removed and rinsed in double distilled water. Tissue was then dehydrated in 70% ethanol for 3 minutes, 95% ethanol for 2 minutes twice, 100% ethanol for 2 minutes four times, and finally cleared in Xylene for 2 minutes four times. The slides were cover slipped with mounting media (Biocare, EM897L) and left to dry.

For analysis and quantification of staining, slides were scanned on Zeiss Axio Scan.Z1. Analysis was conducted on the software Halo V4.0. Cells were segmented by a trained segmentation algorithm. Brightfield analysis on Halo used color deconvolution to separate chromogenic stains. A reference cell was selected as a stain standard, and real time tuning was used to quality control stain deconvolution. Finally, cellular stain optical density was determined for each cell, defined as pixel color intensity across total cell area for each cell analyzed. Increased optical density reflected increased intensity of staining and was quantified for comparison between treatment conditions.

*Target Inhibition by Jess Automated Western Blot System*

Inhibition of PARP was demonstrated by the relative expression of cleaved PARP. Tumoroids in all treatment conditions (no treatment, PARPi only, radiation only, combination PARPi and radiation treatment) underwent protein extraction using RIPA buffer and manual vortex agitation. For quantitative immunoassay, the capillary-based Simple Western assay was performed on the Jess system (ProteinSimple, #004-650) using relevant modules (#DM-001 and #SM-FL004, ProteinSimple) according to the manufacturer’s protocol. For each condition, a protein sample of 2 μg was used per lane. Antibodies for the protein of interest and a loading control protein were run simultaneously. Protein normalization was performed using a relevant module (#DM-PN02, ProteinSimple) according to the manufacturer’s protocol. The Compass software v6.3.0 (ProteinSimple) was used for normalized protein quantification, size determination, and blot construction. Protein expression for each treatment condition was compared to its no treatment condition to quantify relative expression.
