## Supplemental Tables for "Leveraging a Patient-Derived Tumoroid Platform for Precision Radiotherapy: Uncovering DNA Damage Repair Inhibitor-Mediated Radiosensitization and Therapeutic Resistance in Rectal Cancer"

**Supplementary Tables**

| Drug | Concentration | Company | Catalogue Number |
| --- | --- | --- | --- |
| ATMi (KU-55933) | 0.75 μM | Selleckchem | S1092 |
| DNA-PKi:  Nedisertib (M3841) | 0.25 μM | Selleckchem | S8586 |
| ATRi:  (Berzosertib VE-822) | 0.05 μM | Selleckchem | S7102 |
| PARPi:  Veliparib (ABT-888) | 5 μM | Selleckchem | S1004 |
| 5-Fluorouracil | 0.5 μM | Sigma-Aldrich | F6627 |

**Supplementary Table 1**. Summary of DNA Damage Repair Inhibitors

| Antibody | Clone | Catalogue Number | Company | Isotype |
| --- | --- | --- | --- | --- |
| Anti-DNA-PKcs | 3H6 | MA5-15813 | Thermo Fisher | Mouse IgG1 |
| Anti-phospho-DNA-PKcs | polyclonal | ab18192 | Abcam | Rabbit |
| Anti-KAP1 | EPR5216 | ab109287 | Abcam | Rabbit |
| Anti-phospho-KAP1 | polyclonal | PLA0140-100UL | Sigma | Rabbit |
| Anti-CHK1 | 2G1D5 | 2360 | CST | Mouse IgG1 |
| Anti-phospho-CHK1 | 133D3 | 2348 | CST | Rabbit IgG |
| H2A.X | JBW301 | 05-636-1 | EMD Millipore | Mouse IgG1k |
| Anti-PARP1 | E102 | ab32138 | Abcam | Rabbit |
| Anti-cPARP1 | E51 | ab32064 | Abcam | Rabbit |

**Supplementary Table 2**. Antibody Summary for Target Inhibition Validation

| **Tumoroid** | **Intrinsic Radiosensitivity**  **(1-β)** |
| --- | --- |
| RC-MSK-004T | 10% |
| RC-MSK-023T | 11% |
| RC-MSK-042T1 | 15% |
| RC-MSK-042T2 | 16% |
| RC-MSK-012VR | 17% |
| RC-MSK-012PR | 17% |
| RC-MSK-072LM | 17% |
| RC-MSK-012T | 18% |
| RC-MSK-037T2 | 19% |
| RC-MSK-012SP | 22% |
| RC-MSK-022T | 23% |
| RC-MSK-053T2 | 25% |
| RC-MSK-034T1 | 26% |
| RC-MSK-008T | 26% |
| RC-MSK-039T | 28% |

**Supplementary Table 3**: The percent decrease in cell viability with DMSO alone for each tumoroid is shown in order of increasing radiation sensitivity.

| **Tumoroid** | **DNA-PKi** | **ATMi** | **PARPi** | **5-FU** | **ATRi** |
| --- | --- | --- | --- | --- | --- |
| RC-MSK-004T | 1.65 | 1.25 | 1.58 | 1.34 | 1.28 |
| RC-MSK-023T | 1.41 | 1.42 | 1.27 | 0.95 | 1.16 |
| RC-MSK-042T1 | 1.66 | 1.51 | 1.50 | 1.03 | 0.99 |
| RC-MSK-042T2 | 1.36 | 1.31 | 1.53 | 1.18 | 1.16 |
| RC-MSK-072LM | 1.77 | 1.92 | 1.07 | 1.02 | 0.88 |
| RC-MSK-012T | 1.34 | 1.29 | 1.23 | 0.97 | 1.04 |
| RC-MSK-037T2 | 1.55 | 1.51 | 1.26 | 1.19 | 0.97 |
| RC-MSK-022T | 1.40 | 1.28 | 1.27 | 1.12 | 1.11 |
| RC-MSK-053T2 | 1.28 | 1.42 | 0.99 | 0.71 | 0.82 |
| RC-MSK-034T1 | 1.22 | 1.18 | 1.12 | 1.00 | 0.84 |
| RC-MSK-008T | 1.07 | 0.94 | 0.85 | 0.77 | 0.85 |
| RC-MSK-039T | 1.29 | 1.36 | 1.10 | 0.93 | 1.09 |

**Supplementary Table 4**: Slope ratios for each tumoroid in each drug condition relative to DMSO are shown. The degree of sensitization to radiation by drug treatment is characterized by slope ratios comparing the slope of each drug condition to the slope of DMSO. A slope ratio greater than 1 indicates sensitization by drug. A slope ratio less than 1 indicates a lack of sensitization.
